# Hierarchy of immunogenicity of *Plasmodium falciparum* merozoite antigens and vaccine candidates in first human infection

**DOI:** 10.64898/2026.10.18.26365849

**Authors:** Joshua Waterhouse, D Herbert Opi, Linda Reiling, Lee Yeoh, Kaitlin Pekin, Eizo Takshima, Adam Thomas, Hikaru Nagaoka, James S McCarthy, Christian R Engwerda, Bridget E Barber, Michelle J Boyle, James G Beeson

## Abstract

**Background:** Merozoite antigens are targets of acquired immunity and are promising targets for next-generation malaria vaccines and serosurveillance tools. However, the immunogenicity of each antigen and their induction of antibodies following infection remains unclear, impacting vaccine design, antigen selection in serosurveillance, and our understanding of naturally-acquired immunity.

**Methods:** To address this, we assessed IgG and IgM responses to whole merozoites and individual merozoite antigens in 81 malaria-naïve adults who experienced a blood-stage *P. falciparum* Controlled Human Malaria Infection (CHMI).

**Results:** Significant acquisition of IgG and IgM against whole merozoites could be detected as early as two weeks after infection. For IgG responses, a strong hierarchy of immunogenicity among merozoite antigens was observed, with several prominent antigen targets. In contrast, IgM was acquired to most antigens and was less hierarchical with a broader and less antigen-dominant acquisition profile. Participants clustered into distinct immune-acquisition groups according to the breadth and magnitude of their antibody responses, suggesting that individual variation in the development of malaria immunity begins as early as first exposure.

**Conclusions:** Our findings provide important insights into the immunogenicity of merozoite antigens under consideration for vaccine development and identify antigens that could be used in the development of malaria sero-surveillance tools.

## Introduction

Malaria remains a major global health burden, responsible for over 600,000 deaths each year [1]. In endemic regions, individuals gradually develop immunity to malaria over multiple infections, with some children able to acquire protection against severe symptoms after only a few exposures [2–4]. Antibodies to merozoites, the life-stage of the parasite responsible for erythrocyte invasion, have been strongly linked to naturally-acquired protection from malaria [5–10]. These antibodies can suppress blood-stage parasite replication by inhibiting erythrocyte invasion and recruiting other components of the immune system via their Fc fragment, mediating complement fixation and activation, opsonic phagocytosis and cellular cytotoxicity [11–13]. Consequently, merozoites are attractive targets for next-generation malaria vaccines.

Understanding antibody acquisition following infection can provide important insight into antigen immunogenicity and immunodominance. This can help inform antigen selection for vaccine development, since antigen immunogenicity is essential for vaccine efficacy, and an antigen that is immunogenic with infection is highly likely to be immunogenic when used in vaccines. Given the challenge of poor immune responses to malaria vaccines in some endemic populations [14, 15], selecting highly immunogenic antigens may be an important consideration. Short durability of vaccine efficacy and the requirement for annual boosters is a further challenge [14, 16], and focusing on antigens targeted by antibodies boosted following natural exposure may be an effective means to improve vaccine longevity [16]. Additionally, because merozoites are prominent targets of acquired immunity, quantifying antibodies to merozoite antigens has potential for developing malaria serosurveillance tools [7] to identify populations at risk and transmission hotspots or monitor for transmission in near-elimination and post-elimination settings.

Antibody responses to first malaria infection may be particularly useful in understanding antigen immunogenicity. Studies of acquired immunity frequently focus on adults or older children in malaria-endemic settings with complex or unknown exposure histories, including exposure to genetically diverse *Plasmodium* species, making it difficult to define the temporal pattern of antibody acquisition with successive malaria infections [7, 8, 17, 18]. Antibodies elicited by first infection reflect the natural immunogenicity of antigens without prior immune priming. These antibodies may influence subsequent responses to infections or vaccines, and may be markers of immunologic imprinting, making them an important research focus. Children living in areas with moderate-to-high transmission often experience their first malaria infection during their first year of life [19–21]. Identifying major targets of first infection antibodies can be used to design vaccines that mitigate potential interference or leverage infection-induced immune boosting to improve efficacy. In addition, knowledge on the immunogenicity of antigens and antibody acquisition after a single infection is valuable for informing the development of sero-surveillance tools for malaria, which have an important role in malaria surveillance in low transmission and pre- and post-elimination settings [22–24].

Controlled human malaria infection (CHMI), in which adult volunteers are experimentally infected with *Plasmodium* parasites in a highly controlled environment, provides unique opportunities for studying the immune response to first malaria infection [25], with specific advantages over field studies. CHMI studies involving malaria-naïve adults from non-endemic countries eliminate the confounding effect of prior exposure and pre-existing immunity, and so offer a chance to observe the initial formation of malaria immune responses. In addition, these studies involve a tightly-controlled infection that reduces variation between participants in their infection duration, parasite burden, peak parasitaemia, and symptoms, all of which can impact antibody responses [26–30]. CHMI can be performed with blood-stage parasites or sporozoites (delivered via mosquito bite or injection), allowing for further control of exposure.

Antibody responses to merozoites following first CHMI remain poorly defined, particularly the identity of major antigen targets. In previous CHMI studies, it has been shown that a single *P. falciparum* infection with low peak parasitemia is sufficient to induce antibodies to merozoites [31]. To date, only a limited number of antigens have been investigated as antibody targets in CHMI studies, and there is little data directly comparing antibody acquisition between antigens. Among the antibody targets best characterised in CHMI studies are merozoite antigens MSP1_19_ and AMA1 [32–40], and sporozoite antigens CSP and TRAP [33, 41, 42]. A small number of studies have assessed antibodies to other merozoite antigens, including surface proteins (MSP2, MSP4, MSP5, MSP6) and antigens released from the micronemes (EBA175, EBA181) or rhoptries (Rh2, Rh5) [13, 29, 37, 43–45]. In addition, studies of antibody responses following CHMI have predominantly focused on IgG responses, with very little data on other merozoite antibody isotypes such as IgM [13, 44, 46], which can play an important role in immunity.

To identify prominent merozoite targets of first infection antibodies, we assessed malaria-naïve adults (n = 81) following their first blood-stage CHMI for antibodies to whole intact merozoites and a panel of merozoite antigens (n = 31). Antigens were selected based on their known exposure on the merozoite surface before or during erythrocyte invasion, being targets of acquired antibodies in malaria-endemic populations, and, for some antigens, having a demonstrated role in erythrocyte invasion or association with naturally-acquired immunity to malaria. Antibodies were quantified at baseline (pre-infection), two weeks post-infection, and at the study end point (day 28-45). We examined the acquisition of IgG and IgM, identifying the most immunogenic antigens, evaluated the co-acquisition of antibodies to different antigens, and revealed how responses to antigens varied among study participants.

## Results

### First blood-stage infection induced IgG and IgM against whole merozoites

To investigate merozoite antibodies induced by a single *P. falciparum* infection, we assessed IgG and IgM levels in malaria-naïve adults following their first blood-stage CHMI. Samples were collected from 81 adult participants across 17 different CHMI trials (Supplementary Materials**, Table S1**); 76/81 (77%) were male, and the median age was 24 years. Participants were intravenously inoculated with ∼2,800 *P. falciparum*-infected erythrocytes, then administered antimalarial drugs once parasitaemia reached ∼20,000 parasite per mL blood (**Fig. 1A**). Plasma samples (n = 81, 3 timepoints) were collected from pre-infection (day 0), two-weeks post-infection (days 14-16), and at end-of-study (EOS, days 28-45).

**Figure 1.**
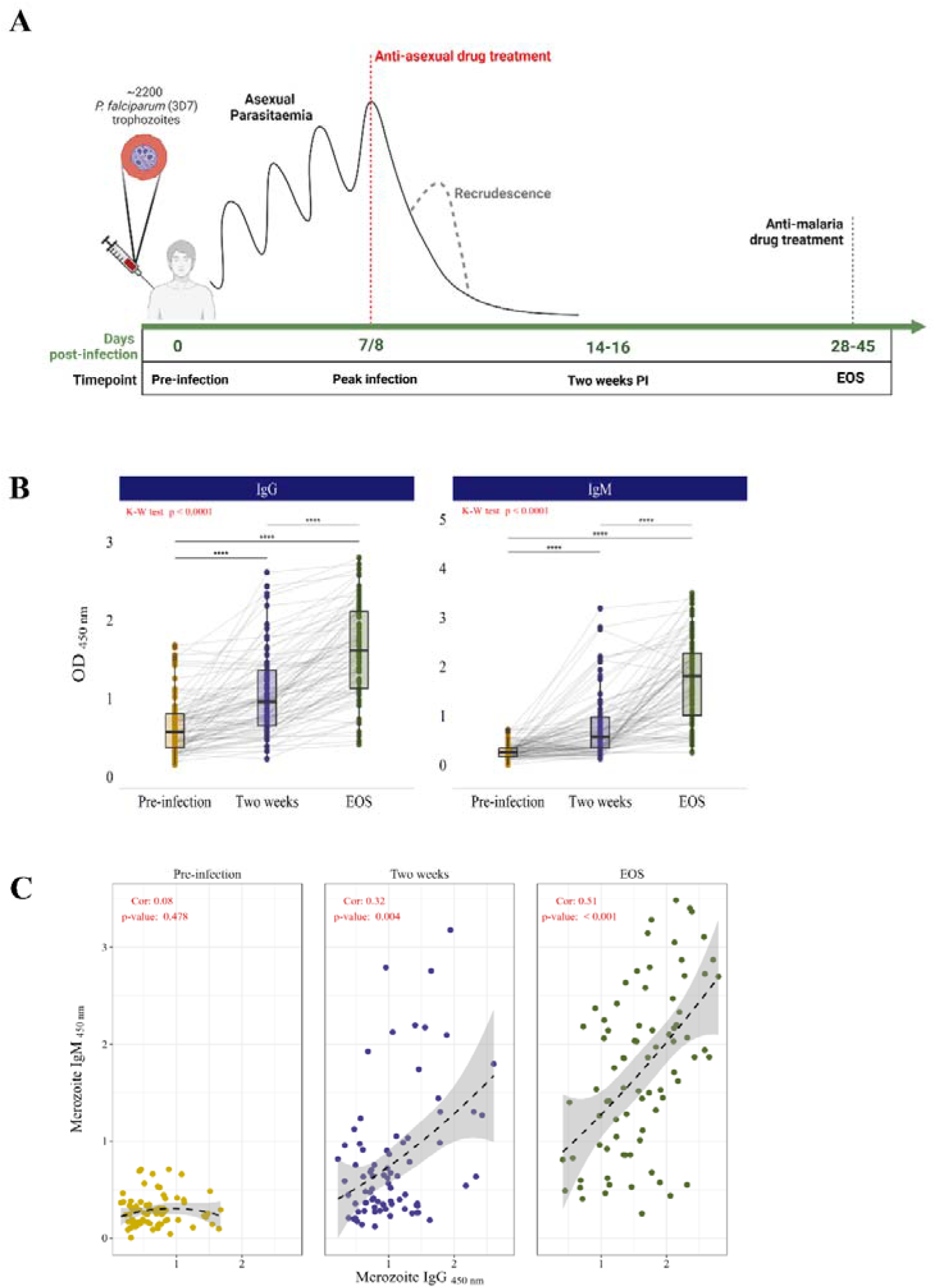
First *P. falciparum* infection induced antibodies against the whole merozoite surface. (A) Schematic of blood-stage CHMI trials. Malaria-naive adults (n = 81) were inoculated with ∼ 2800 *P. falciparum*-parasitised RBCs at day 0. Asexual parasitaemia was monitored daily and peaked at 7-8 days post-infection (dpi) with ∼20,000 parasites per mL blood, at which time antimalarial drugs were administered. Recrudescence was treated using a course of artemether-lumefantrine. At end-of-study (EOS) all participants received another course of antimalarial drugs to clear any remaining parasites. Plasma samples collected at 0 dpi (’Pre-infection’), 14-16 dpi (’Two weeks PI’), and 28-45 dpi (end-of-study, ‘EOS’) were tested for merozoite antibodies. (B) Whole merozoite antibodies measured by ELISA (OD_450 nm_), including IgG (left) and IgM (right), for samples from pre-infection, two-weeks PI, and EOS. Grey lines represent participant samples over time. Antibody levels between time-points compared using Kruskal-Wallis test paired with Dunn’s test. Significant Dunn’s test outcomes represented by stars. (C) Spearman’s correlations between IgG and IgM to the whole merozoite surface between time-points. Spearman correlation coefficients and p-values shown.

We first quantified IgG and IgM reactivity against whole *P. falciparum* merozoites among participants before and after infection. Most participants tested seropositive for IgG (61%, 49/81) and IgM (87%, 70/81) to the whole merozoite surface by EOS (**Fig. 1B**). IgG and IgM reactivity to the merozoite surface at EOS were moderately correlated (rho: 0.51, p < 0.001; **Fig. 1C**). Kinetics of IgG and IgM development varied considerably between individuals, with some having detectable antibodies by two-weeks post-infection, whereas others developed an antibody response between two-weeks and EOS (**Fig. 1B**).

### Antibodies elicited by first infection target a range of merozoite surface antigens

We next aimed to identify merozoite antigens targeted by acquired antibodies, and to determine whether first infection induced antibodies against antigens involved in erythrocyte invasion, or targets of antibodies linked to malaria immunity [5, 7, 8, 47–49]. Antigens included in the panel are expressed by merozoites and known to be exposed on the merozoite surface before erythrocyte invasion is complete, meaning they could be feasible targets of protective antibodies. The panel included many antigens that have been considered as merozoite vaccine candidates. CSP and Pfs230, expressed by sporozoites and gametocytes, respectively, were included to test whether a single blood-stage infection elicits antibodies against vaccine-relevant antigens expressed by other *P. falciparum* life-stages. Although exposure to sporozoites does not occur in induced blood-stage CHMI studies, gametocytemia can develop with gametocytes in the peripheral blood detectable by PCR at 10-12 days after infection [50]. Antibodies to Pfs230 have been reported to develop through natural exposure [51, 52].

We found that CHMI participants acquired IgG to a range of antigens expressed on the merozoite surface (**Fig. 2**). There was strong evidence (p < 0.001) that participants acquired IgG to well-established targets of acquired immunity, such as MSP1_19_, MSP2 and AMA1, and multiple antigens that have been less well studied, especially in CHMI studies, including MSP3, MSP6, and P12. There was also significant acquisition of an IgG response to proteins in the Rh5-invasion complex, which are in clinical trials and vaccine development (collectively these are known as the PCRCR proteins and include Rh5, CyRPA, Ripr, CSS, PTRAMP). In all, there was significant acquisition of IgG for 22 merozoite antigens. There was no significant increase in participant IgG levels from baseline to EOS for MSP4, P41, EBA140 (RIII-V), EBA175 (RII), CyRPA, Rh2, Rh4, and RALP1. Of note, there was also no significant change in response to CSP or Pfs230.

**Figure 2.**
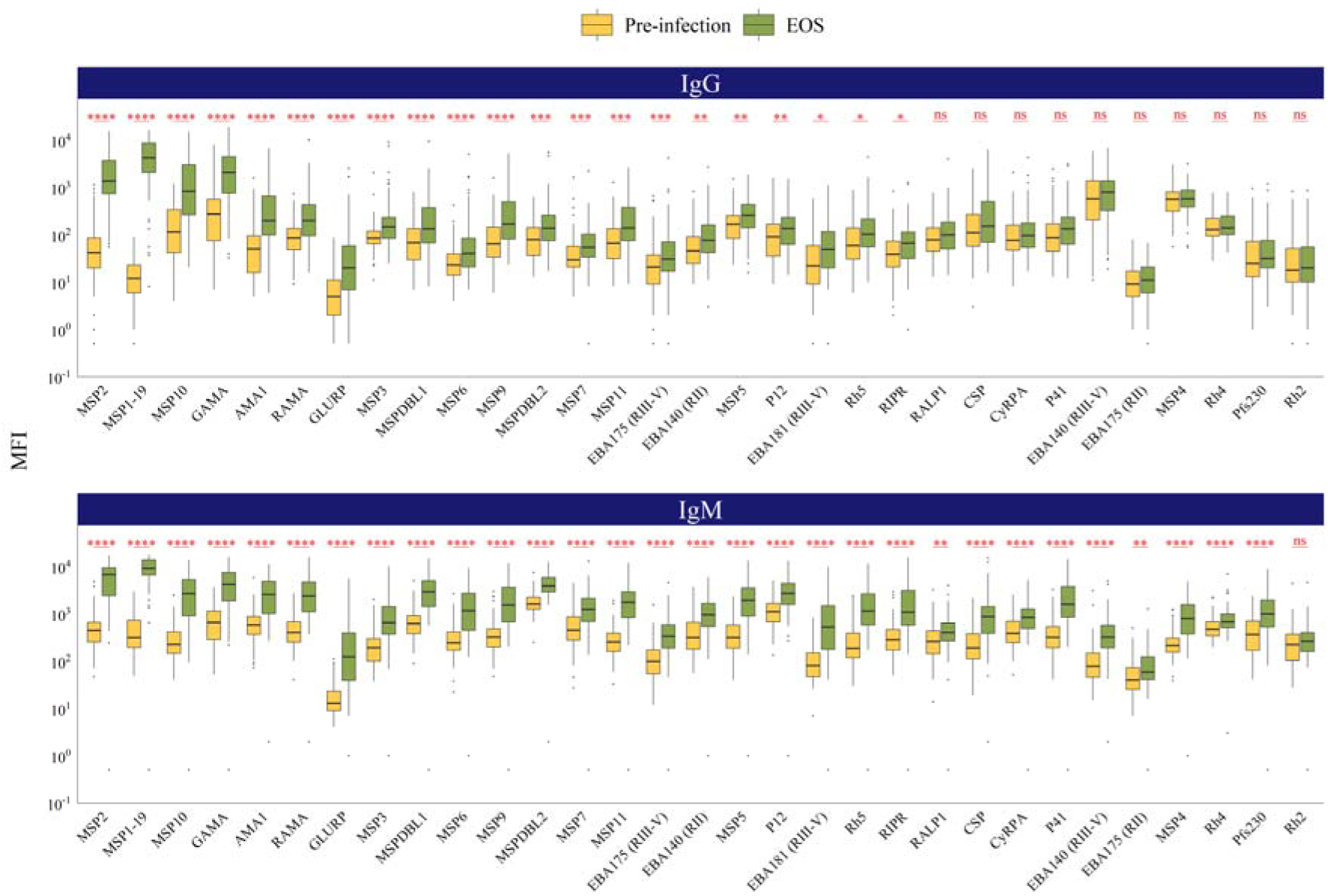
First CHMI induces antibodies to a range of merozoite antigens. Acquisition of IgG and IgM to *P. falciparum* antigens. Antibody levels compared between pre-infection (gold) and EOS (green) using a paired Wilcoxon signed-rank test. Significant acquisition of antibodies reported as *p<0.05, **p<0.01, ***p<0.001, ****p<0.0001, or ns for non-significant (p≥0.05).

CHMI participants also had a broad IgM response, with significant acquisition of IgM to all assessed antigens except Rh2. Interestingly, this included antigens where a poor IgG response was observed. There was significant IgM reactivity to CSP and Pfs230. While CSP is not expressed by blood-stage *P. falciparum*, in prior studies cross-reactive antibody responses between CSP and blood-stage antigens have been reported due to shared or similar epitopes [53].

### A subgroup of merozoite surface antigens are prominent targets of IgG acquired following first infection

To assess whether there is an hierarchy of immunogenicity among antigens, we compared antigen acquisition across antigens by seroprevalence and reactivity fold-change (EOS_MFI_ / Pre-infection_MFI_) (**Fig. 3**). This demonstrated that there is a hierarchy of antibody responses with a subset of antigens that are most immunogenic, generating both the highest fold-change increase in specific IgG and higher seroprevalence across the group. MSP1_19_ was the most prominent IgG target, acquired by most participants (95%, 77/81) and with the greatest median reactivity fold-change (median IgG log_2_ fold-change (FC): 179) (**Fig. 3**). MSP2 was also a prominent IgG target with high seroprevalence by EOS (89%, 72/81) and higher reactivity fold-change than most other targets (FC: 34).

**Figure 3.**
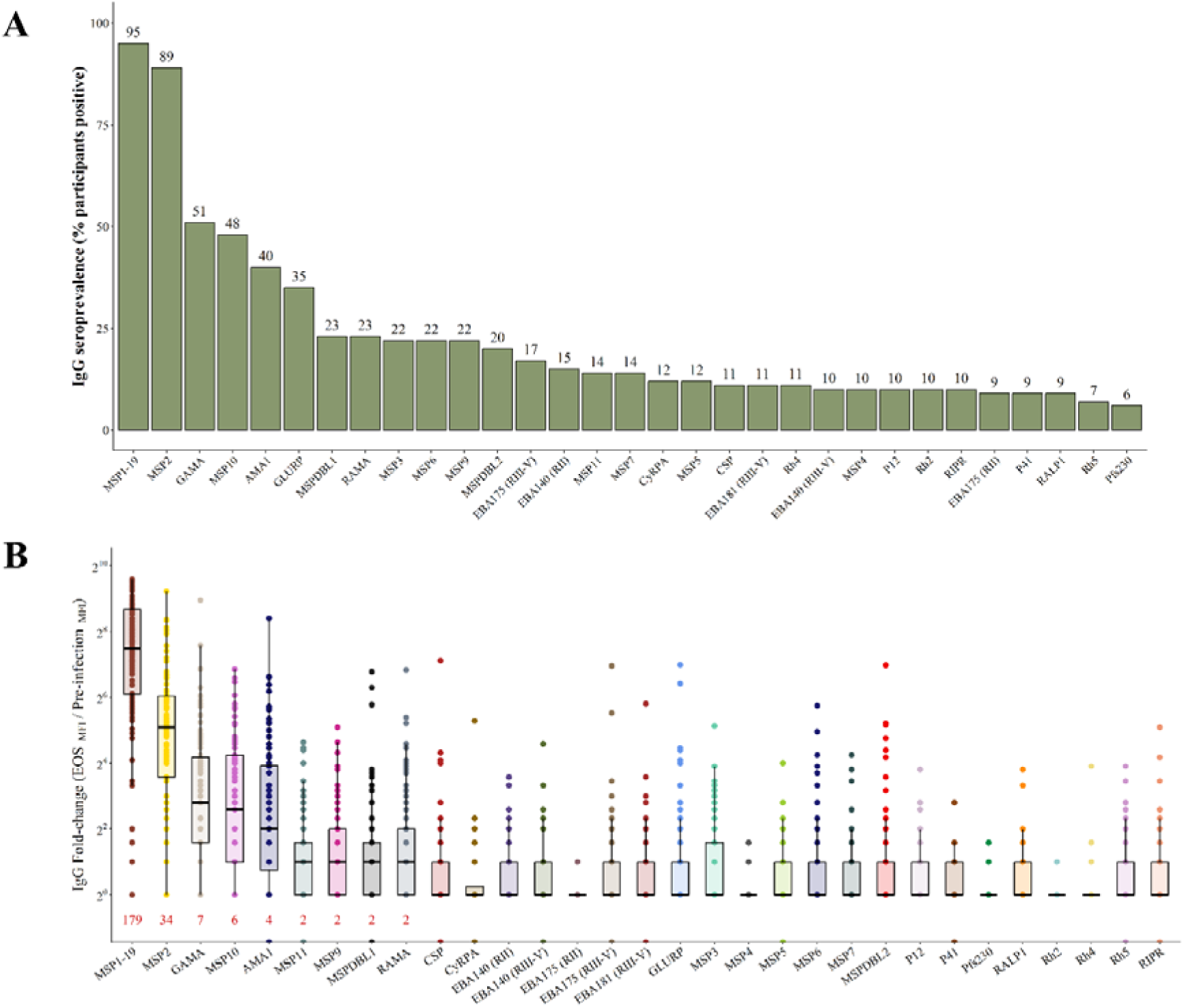
IgG seroprevalence and reactivity fold-change across antigens. (A) Antigen IgG seroprevalence (% of participants seropositive) at EOS. Antigens ordered high-to-low by seroprevalence. (B) Antigen reactivity fold-change between pre-infection and EOS. Median fold-change above 1 shown in red text under each boxplot, and reported for top five most reactive antigens.

Approximately half the participants acquired IgG to GAMA (51%, 41/81) and MSP10 (48%, 39/81), with median fold-change in reactivity substantially higher than most panel antigens (GAMA FC: 7, MSP10 FC: 6), antigens for which there is a limited understanding as targets of immunity. AMA1 and GLURP were notable targets of IgG, acquired by 40% (24/81) and 35% (28/81) of participants, respectively, and with a higher fold-change than most other antigens. For other antigens, IgG was infrequently acquired (< 25% of participants), with relatively low median IgG fold-change for RAMA, MSPDBL1, MSP9, and MSP11; responses were very low for the remaining antigens. Overall, the seroprevalence of IgG responses to 20 antigens matched or exceeded 10% by EOS. These findings suggest that many antigens are less immunogenic, and are less prominent targets of acquired IgG, and that multiple infections, or infections with greater parasitaemia or duration, may be required for the acquisition of specific IgG to these targets.

### Broad IgM reactivity with lower antigen-specific skewing relative to IgG

In contrast, the pattern of antigen-specific IgM responses and fold-change was very different to IgG; 15 antigens had IgM prevalence of >50% and there was a substantial fold-change for most antigens. The profile of seroprevalence across antigens for IgM was significantly different to IgG (p<0.0001). Similar to IgG, MSP1_19_ was the most prominent target of IgM responses; this was acquired by nearly all participants (94%, 79/81). Likewise, the highest IgM reactivity fold-change (FC) was seen for this antigen (FC: 20.3) (**Fig. 4**). MSP2, MSP10, and MSP11 were also major IgM targets, acquired by most participants (78%, 78%, and 79%, respectively) and with notable reactivity fold-change (FC: 12.1, FC: 9.7, and FC: 5.8, respectively). By EOS, an increase in median IgM levels across participants was detected (FC > 1) for most antigens, in contrast to IgG where similar increase was seen for only a few targets. Also in contrast to IgG, the highest medium IgM fold-change values (e.g. MSP1_19_, FC=20.3; MSP2, FC=12.1) were substantially lower than the highest IgG fold change values observed (E.g. MSP1_19_ FC=179, MSP2, FC=34). These trends suggest the IgM response to first infection is more diffuse across a range of antigens and less hierarchical, in contrast to IgG which revealed a strong hierarchy of responses across the different antigens. Notably, there were significant IgM responses after a single infection to Rh5 and Ripr, antigens which are both currently in vaccine trials. However, the generally low IgG response to these antigens despite higher prevalence of IgM reactivity suggests class switching has not occurred, and that further exposure (infections or parasite load) may be needed for greater isotype switching from IgM to IgG.

**Figure 4.**
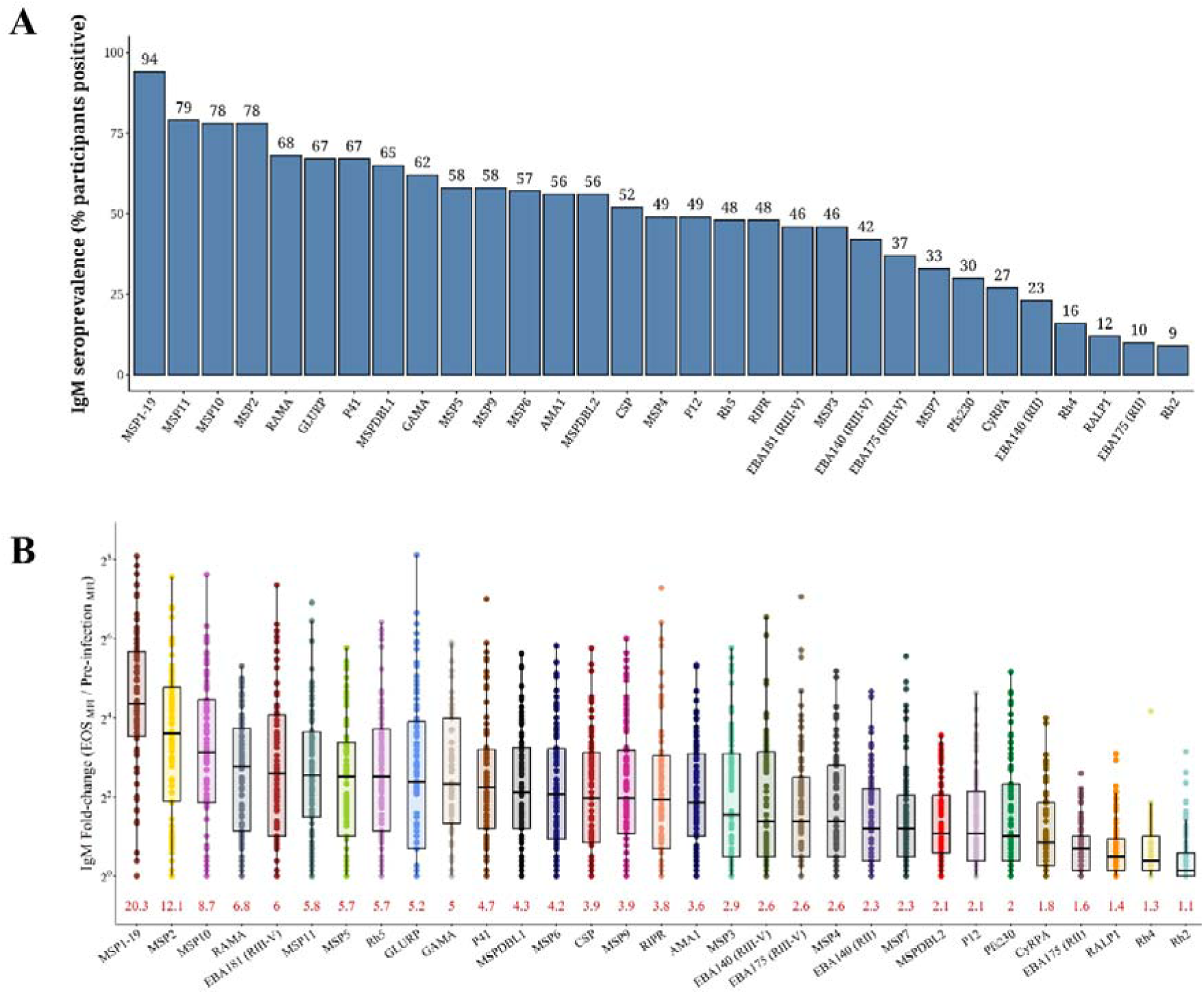
IgM seroprevalence and reactivity fold-change across antigens. (A) Antigen IgM seroprevalence (% of participants seropositive) at EOS. Antigens ordered high-to-low by seroprevalence. (B) Antigen reactivity fold-change between pre-infection and EOS. Median fold-change above 1 shown in red text under each boxplot, and reported for top five most reactive antigens. Seroprevalence across antigens for IgM was significantly different to IgG (p<0.0001).

### Divergent patterns in IgG and IgM co-acquisition across merozoite antigens

We next investigated the co-acquisition of antigen antibodies by performing Spearman’s rank-sum correlations and ordering antigens via hierarchical clustering. Correlation strength was classified as either weak (|rho| < 0.5), moderate (0.5 ≤ |rho| < 0.7), or strong (|rho| ≥ 0.7); only significant correlations (p < 0.05) are shown (**Fig. 5**).

**Figure 5.**
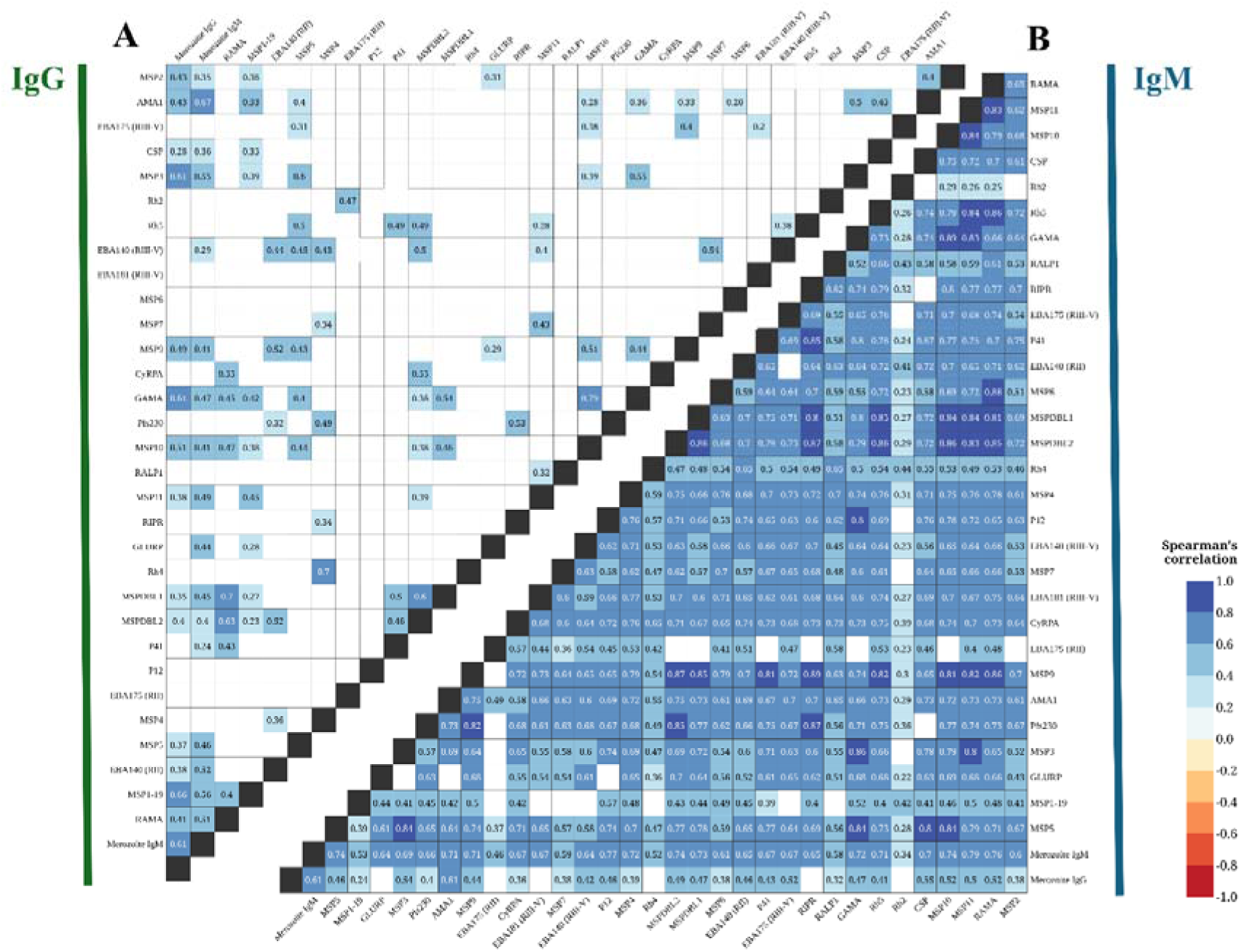
Correlation heatmap of merozoite antigen-specific antibodies. Pairwise correlations between antigen-specific IgG (A) and IgM (B) at EOS. Correlations assessed using Spearman’s rank-sum correlation. Rho values displayed in cells with shading by correlation intensity and direction (blue = positive, red = negative). Non-significant correlations (p ≥ 0.05) left blank.

The strongest correlation of IgG levels between antigens was observed between GAMA and MSP10 (rho = 0.79) (**Fig. 5A**). In addition, there were moderate-to-strong IgG correlations for a trio of antigens: RAMA, MSPDBL1, MSPDBL2. For other antigens, IgG correlations were either moderate, weak, or non-significant, suggesting a weak relationship in the IgG response to these targets following first infection. In contrast, IgM to most antigens had moderate-to-strong correlations (**Fig. 5B**), and these broad antibody correlations suggest that antigen IgM levels are influenced by an individual’s overall IgM response, in conjunction with antigen-specific immunogenicity.

### Different merozoite antibody response profiles among CHMI participants

To identify potential patterns in antibody acquisition across individuals, CHMI participants were clustered based on their merozoite antibody levels at EOS, with resulting clusters evaluated via principal component analysis (PCA). Three distinct groups emerged with clear separation (**Fig. 6**). These groups differed significantly in antibody breadth and magnitude. IgG and IgM responses were highest in cluster 2 (n=26), moderate in cluster 1 (n=40), and generally low in cluster 3 (n=15). Cluster 2 participants also exhibited the highest and broadest antibody responses to individual antigens, whereas participants in cluster 1 showed more moderate responses to a subset of antigens, and those in cluster 3 showed relatively low or negligible antibody responses to first infection (**Fig. 6B-C**, **Fig. 7, Supplementary Fig S2**). Differences between the three clusters were significant for most antigens in the array. For IgG responses, a group of antigens had the most prominent and consistent responses across study participants; these include MSP1_19_, MSP2, GAMA, and MSP10 (**Fig 7**). A second set of antigens were also prominent targets, mainly in cluster 2, including AMA1, MSP9, RAMA, MSPDBL1 and MSPDBL2. Responses to other antigens were much more variable among participants. Taken together, these trends highlight the individual variation in the merozoite antibody response to first infection, suggesting that individual variation in malaria immunity development begins as early as the first exposure, and highlights a hierarchy of immunogenicity of antigens for IgG responses.

**Figure 6.**
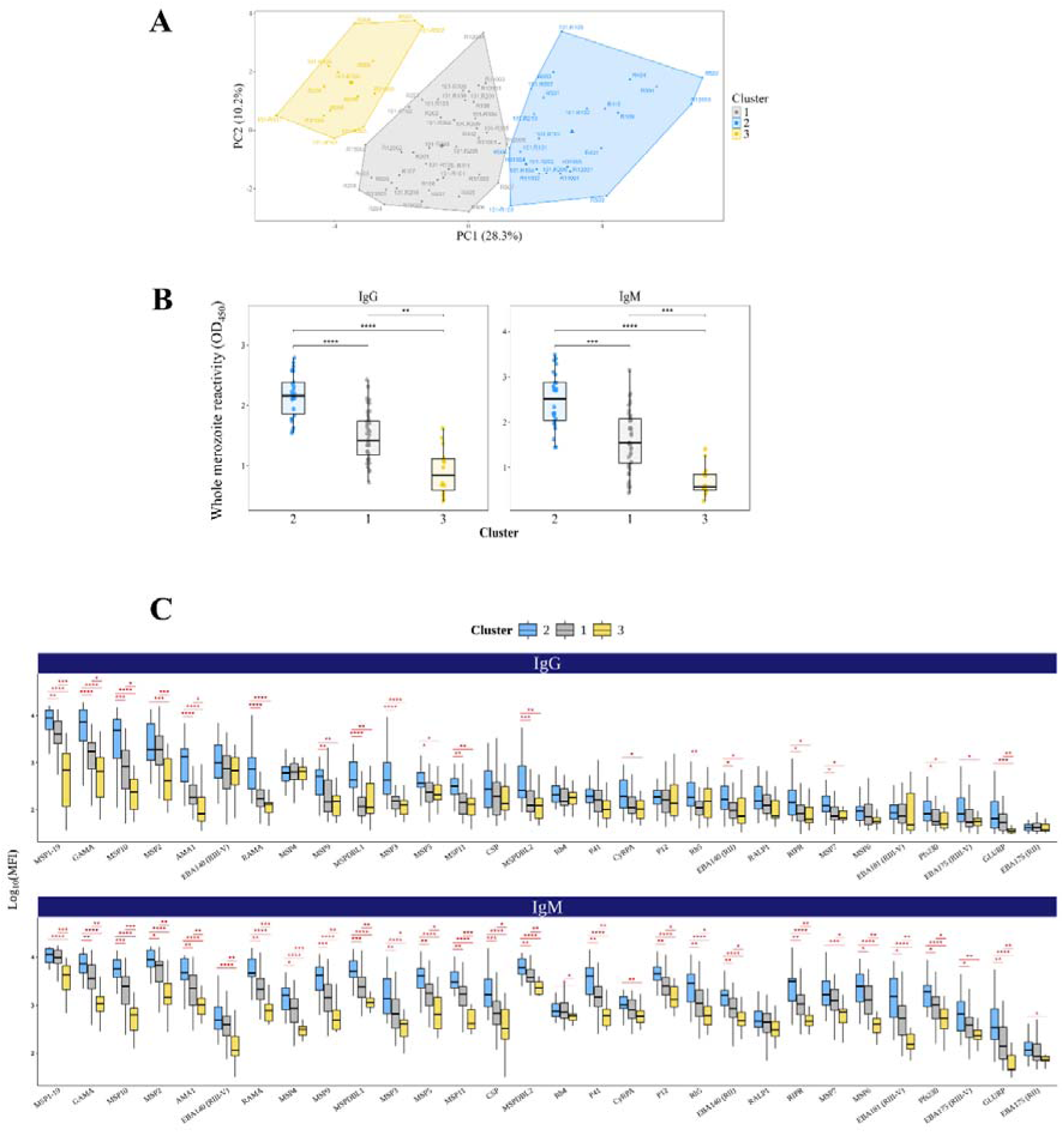
CHMI participants cluster by merozoite antibody responses. (A) Principal component analysis (PCA) of CHMI participants based on merozoite antigen-specific IgG responses. Principal components 1 and 2 (PC1 and PC2) explain 28.3% and 10.2% of total variation, respectively. Participants coloured by clusters identified via k-means clustering (k = 3). (B) IgG (left) and IgM (right) levels to the whole merozoite surface compared between participant clusters. Clusters ordered by median reactivity. (C) Antigen-specific merozoite antigen antibody levels between participant clusters. IgG (top) and IgM (below) levels to merozoite antigens compared between clusters 2 (blue), 1 (grey), and 3 (gold). All group comparisons made using a Kruskal-Wallis test paired with Dunn’s test. Significant Dunn’s test outcomes reported as: *p<0.05; **p<0.01; ***p<0.001; ****p<0.0001. Non-significant correlations (p ≥ 0.05) left blank.

**Figure 7.**
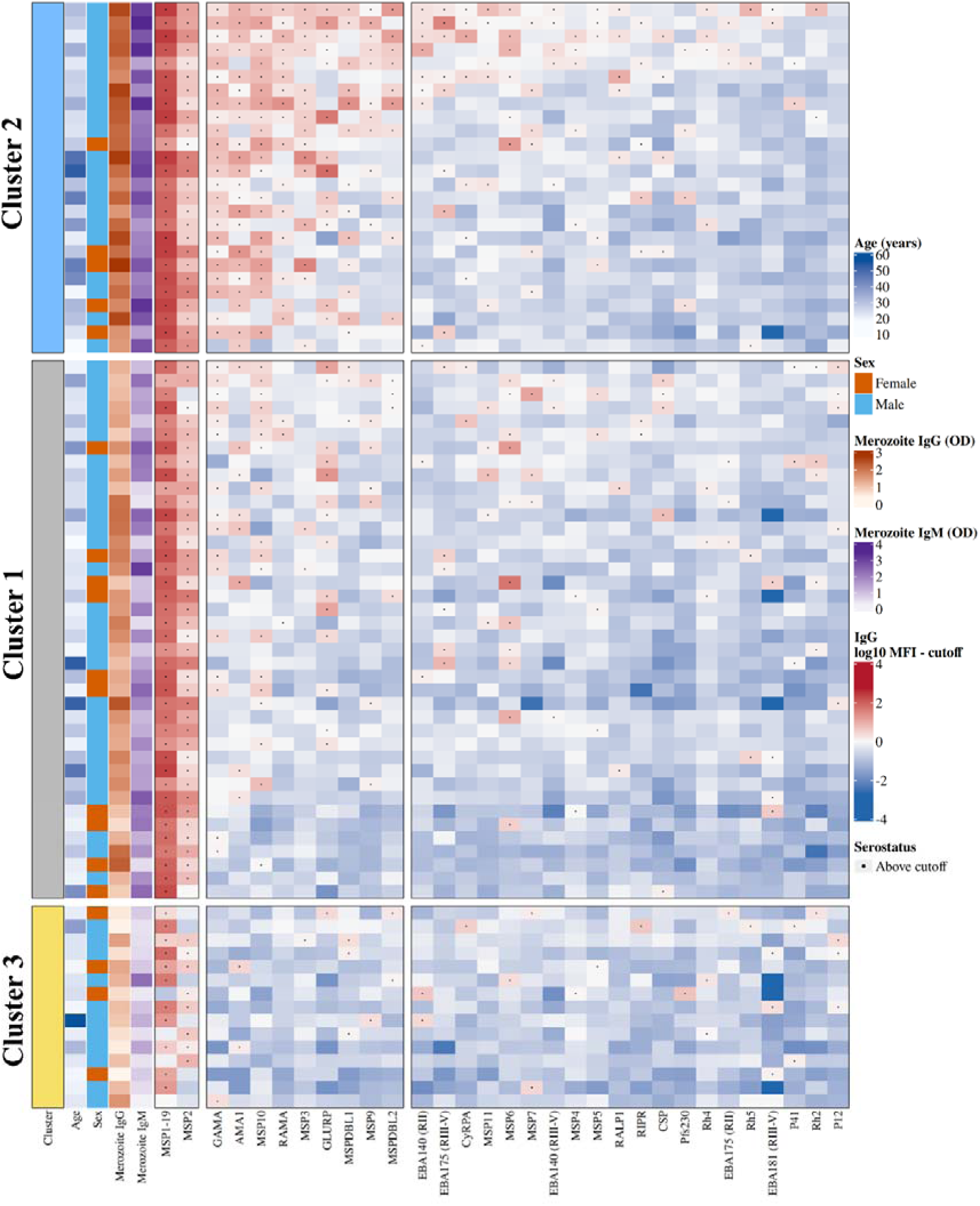
Heatmap of merozoite antibodies show distinct reactivity profiles between participant clusters. Antigen-specific IgG reactivity at end of study (EOS) is shown for participants (rows) and antigens (columns). Cell colours represent log10-transformed MFI relative to antigen-specific seropositivity cutoffs (defined as the 95th percentile of log10-transformed MFI at pre-infection). Cutoffs were subtracted from EOS values, with red indicating reactivity above the cutoff, white at the cutoff, and blue below the cutoff. Black dots indicate seropositivity. Annotations (left) include participant age, sex, and whole merozoite IgG and IgM reactivity measured by ELISA. Participants split by pre-determined clusters. Antigens in order according to cluster 2 IgG seroprevalence.

Cluster 2 participants were significantly older (median age 28.6 years) than those in cluster 3 (median 22.0 years, p=0.018), and trended higher than those in cluster 1 (median 23.5 years, p=0.095) (**Fig. S3A**). Splitting participants into age tertiles (Youngest median age; all groups n=27), age was associated with higher IgM to whole merozoites and antibodies to multiple antigens, including IgG to MSP1_19_, MSP3, and RAMA, and IgM to MSP2, MSP4, and CSP (**Fig. S3B-C**). Similar antibody levels were observed between male and female participants, with similar sex distributions between clusters (**Fig. S3D-E**).

## Discussion

Antibodies targeting merozoites are a key component of naturally-acquired immunity to malaria; however, their acquisition following initial exposure is not fully understood. This knowledge is valuable for informing antigen selection in vaccine development, understanding or predicting boosting effects of natural exposure, and designing sero-surveillance tools for malaria. Here, we provide important new data on the acquisition of IgG and IgM to whole merozoites and individual merozoite antigens. There was significant IgG and IgM acquisition against whole merozoites as early as two weeks after infection. Importantly, our data reveal that there is a hierarchy of immunogenicity among merozoite antigens for IgG responses, and we identified a subset of merozoite antigens that were the most prominent targets of IgG, and antigens for which there was no significant IgG generation. In contrast, IgM was far less hierarchical, with acquisition of IgM to most antigens and with a broader and less antigen-dominant pattern. Participants clustered into distinct immune-acquisition groups according to the breadth and magnitude of their antibody responses, suggesting that individual variation in malaria immunity development begins as early as the first exposure.

In response to first infection, malaria-naïve adults developed antibodies to a range of merozoite antigens. Several of these antigens are well-established as targets of IgG following CHMI, including MSP1_19_, MSP2, GLURP, and AMA1 [13, 29, 32, 34–36, 44, 54], whereas other targets have received little or no previous evaluation as immune targets following CHMI, such as RAMA, MSP11, GAMA, and MSP10. Furthermore, the IgM response to CHMI has been little studied, with IgM to only a handful of merozoite targets previously assessed [13, 44, 46]. We found that CHMI elicits IgM that recognises most merozoite antigens.

An important finding from this study was the identification of MSP1_19_, MSP2, GAMA, and MSP10 as the most prominent IgG targets following first infection. MSP1 and MSP2 are the most abundant GPI-anchored proteins on the merozoite surface [55], and this high abundance may be an important factor of their immunogenicity, in addition to other factors such as their surface location and presence of T-cell and B-cell epitopes. Other moderately immunogenic antigens included AMA1, MSP9, MSPDBL1, GLURP and RAMA. The prominence of these antigens as targets of IgG following first infection can be used to inform vaccine development in several ways. First, our findings highlight antigen immunogenicity in malaria-naïve people. Malaria vaccines predominantly target young children, a population with little or no prior malaria exposure, and in endemic regions where transmission is expected to decline, vaccines will increasingly be administered to people with little or no pre-existing immunity. Antigen immunogenicity with first infection may reflect the immune reactivity in these groups. Second, the higher immunogenicity of these antigens may indicate that there will be boosting of responses with natural exposure if these antigens were included in vaccines. Supporting this, malaria-naïve adults vaccinated against *P. falciparum* antigens had substantial antibody boosting with subsequent challenge [32] and Ugandan recipients of blood-stage vaccine BK-SE36 had antibody boosting with post-vaccination malaria exposure [56]. In contrast, no boosting of responses by natural exposure appears to occur with the RTS,S vaccine [16, 57].

Our findings are also relevant to developing and implementing sero-surveillance tools that use acquired anti-malarial antibodies to detect or quantify malaria exposure. The most immunogenic antigens may be suitable for use in sero-surveillance strategies to detect malaria exposure in populations situated in low transmission areas, or pre- and post-elimination settings, where high sensitivity is needed to detect infection events that may not be captured through standard case-reporting systems. Our data found that most subjects acquired antibodies to a subset of 3 or 4 antigens (MSP1_19_, MSP2, GAMA and MSP10), suggesting these antigens might be suitable for sero-surveillance assays where high sensitivity is required. It is striking that 98.8% (80/81) of subjects were positive for IgG to MSP1 or MSP2, and there was significant IgG reactivity to these antigens in all participant clusters, suggesting that these antigens would be valuable in sero-surveillance applications to detect exposure in low transmission or near-elimination settings.

Prominent antibody responses to MSP1 and MSP2 following first infection supports the role of these antigens as important targets in the early formation of naturally-acquired immunity. It has been argued that limited malaria exposure, involving three or four infections, may be sufficient to confer strain-transcending malaria immunity [3]. In addition, one or two exposures has been sufficient to confer immunity against severe malaria and blood-stage parasites in some contexts [4, 58–60]. Antibodies potently induced against MSP1 and MSP2 following first infection may play a key role in this protection. GAMA is an essential GPI-anchored micronemal protein, with a putative role in parasite erythrocyte invasion via the sialic-independent pathway [61, 62]. MSP10 is a merozoite surface protein. that complexes with GAMA via its N-terminal region [63–65]. GAMA-MSP10 complexing may be a factor in the strong antibody co-acquisition observed in this study. A growing body of work suggests that GAMA and MSP10 are important targets of the immune response to malaria. Naturally-acquired antibodies to both GAMA and MSP10 have been found to inhibit parasite invasion *in vitro* and linked to protection from malaria in endemic populations [7, 8, 61, 63]. Limited attention has been given to either GAMA or MSP10 in vaccine development, with neither antigen targeted by vaccines that reached clinical trials. GAMA and MSP10 have both been found to be immunogenic in adult travellers following their first clinical malaria episode [66].

While Rh5, Ripr, and CyRPA were not prominent targets of IgG acquired after first CHMI, there was significant induction of IgG to Rh5 and Ripr, which is encouraging since these are currently actively being pursued as vaccine candidates and in clinical trials [67, 68]. Data suggests that antibodies to Rh5, Ripr, and CyRPA are less prominent in naturally-acquired immunity compared to some other antigens [69–75]. However, naturally-acquired IgG to Rh5, Ripr, and CyRPA has been associated with protection from malaria [7, 71, 73] suggesting that antibodies to these antigens can play an important role in immunity. This study suggests that a first, low-parasitemia *P. falciparum* infection infrequently induces IgG to Rh5, Ripr, and CyRPA; however, there was acquisition of specific IgM, suggesting that greater exposure (multiple episodes or higher parasitemia load or duration) may be required for more robust IgG responses.

CHMI potently elicited IgM that recognised numerous merozoite vaccine candidates, including Rh5, MSP2, AMA1, Ripr, CyRPA, MSP1, MSP3 and GLURP. Our findings suggest that first infection can induce a potent IgM response to multiple merozoite vaccine candidate antigens, and further investigation is needed to understand their role in immunity and if these antibodies can impact vaccine outcomes. An association between IgM responses to merozoites and protection from malaria has been reported in some studies [13]. Overall, it was notable that the IgM response was much broader and less hierarchical than the IgG response. The broad multi-antigen response of IgM may help reduce the potential of antigenic polymorphism, well recognised in some of these antigens, and immune escape. Additionally, development of IgM to CSP was unexpected, since participants were exposed to asexual blood-stage *P. falciparum* parasites and CSP is expressed by sporozoites. This could be due to antibody cross-reactivity with similar epitopes shared across antigens [76–78]. IgM was also reactive to Pfs230, which is expressed by gametocytes. PCR-detectable gametocytes in this infection model emerge after the administration of some anti-malarial drugs that do not block gametocytogenesis (∼10 days post-inoculation) [50].

Participants clustered into different response profiles according to the magnitude and breadth of their merozoite antibody response to first infection. Previous CHMI studies have also reported substantial individual variation in merozoite antibody levels, as well as antibody functions, T_FH_ cell composition, inflammation, and adverse events [36, 44, 79]. Our findings extend on these observations by identifying distinct antibody response profiles across individuals, with some exhibiting a robust immune response to a range of antigens, whereas other produced relatively little antibodies to a narrow set of targets. Differences in individual antibody responses may reflect intrinsic differences between individuals such as age and genetics, or the magnitude and duration of parasite exposure, or a combination of both. Multiple clinical factors have been reported to influence antibody responses to CHMI, including cumulative parasite density and latent CMV infection [29, 80]. MHC genotypes were not studied in this cohort, but may also be an important factor. In agreement with previous CHMI studies, merozoite antibody levels positively correlated with age. High individual variation in malaria immunity is often observed in endemic populations [81, 82], and evidence in this study, alongside others, suggests that individual heterogeneity in immune development begins at first infection.

CHMI is a useful model for studying first infection, with some limitations. Participants in this study experienced week-long, low-density infections with parasitaemia reaching ∼20 parasites per microlitre of blood (range typically 5 to 50) [50, 83–86]. By contrast, individuals in endemic regions can experience infections that can persist longer, induce symptoms ranging from mild to severe, and parasitaemia levels that can reach orders of magnitude above those during CHMI [87, 88]. Variation in these infection parameters— duration, symptoms, parasitaemia, as well as parasite genotype—can impact the malaria antibody response [26, 89–92]. However, an advantage of CHMI studies is that participants experience tightly-controlled infection, which minimises exposure variation that can obscure trends in the immune response and better enables analysis of the hierarchy of antibody responses and isotype switching. Age is an important determinant on malaria immunity [93, 94] and, in contrast to participants in this study, individuals living in areas with moderate-to-high transmission are often first exposed to malaria during childhood [4, 19, 20]. In addition, sex has been linked to parasite emergence during CHMI [95], parasite clearance rate with natural infection [96], and malaria vaccine antibody responses and immunity [97, 98]. Further investigation is needed to understand the impacts of age and sex on malaria immunity.

In conclusion, our study revealed a prominent hierarchy in the immunogenicity of merozoite antigens following a single infection in humans, with a strong contrast between IgG and IgM responses in antigen breadth, profile, and dominance. We identified a subset of antigens that were most strongly immunogenic for IgG and found that IgM is generated to most antigens after a single infection. Further, we revealed high heterogeneity in responses among subjects, even when the infectious dose and infection duration was standardised. These findings provide novel insight into antibody responses to first malaria infection, which can be used to inform antigen selection in malaria vaccine design and in developing sero-surveillance tools.

## Materials

### Study cohorts

Controlled human malaria infection (CHMI) studies were conducted following the Induced Blood-Stage Malaria (IBSM) model as previously described [99]. Details of the six studies are included in **Table S1**. In brief, healthy malaria-naïve adults were inoculated with ∼2,800 viable *P. falciparum*-infection RBCs and monitored for symptoms. Peripheral parasitaemia was quantified via qPCR [100]. Participants were administered antimalarial drugs at 7 or 8 days post-inoculation (dpi), when parasitaemia was ∼ 20,000 parasites/mL. Participants with recrudescence of asexual parasites were treated with artemether-lumefantrine. Blood samples were collected in lithium heparin tubes prior to infection (0 dpi), two weeks post-infection (14-16 dpi), and end-of-study (EOS, 28-45 dpi). Plasma was isolated from blood samples via centrifugation, then snap frozen on dry ice and stored at -70 °C. Samples were collected from 81 participants enrolled in 17 independent infection cohorts across six studies. Clinical trials were registered at ClinicalTrials.gov or the Australian and New Zealand Clinical Trials Registry, and originally conducted to test the safety and efficacy of anti-malarial drugs (NCT02867059 tested SJ733 [83]; NCT03542149 tested OZ439 [84]; NCT02783833 tested MMV390048 [85]; ACTRN12620000995976 tested tafenoquine [86]), experimental *P. falciparum* transmission to *Anopheles* mosquitoes (NCT02431637 [50]), or drug gametocytocidal activity and parasite transmission-blocking (NCT02431650 tested OZ439 [50]).

All participants provided written informed consent prior to inclusion. Ethics approval for the collection and use of human samples was obtained from the Human Research and Ethics Committee at QIMR-Berghofer Medical Research Institute (P1479, P3444, P3445) and the Alfred Human Research and Ethics Committee at the Burnet Institute (#225/19).

### Recombinant proteins

Multiplex antibody assays were performed with a panel of 31 P. falciparum recombinant proteins. Most panel proteins are expressed by merozoites and selected based on a known or putative role in merozoite-erythrocyte invasion, or previously found to be targeted by antibodies linked to malaria immunity. Circumsporozoite protein (CSP) and Pfs230— expressed by *P. falciparum* sporozoites and gametocytes, respectively—were included to assess if first blood-stage infection induced antibodies to proteins expressed by non-asexual forms of the parasite. Proteins were expressed using *E. coli*, HEK293 cells, Pichia pastoris (P. pastoris), and Spodoptera frugiperda (Sf21) insect cells, or the Wheat Germ Cell-Free protein expression system. Antigens were produced at multiple institutions, including Burnet Institute (Burnet), the National Institutes of Health (NIH) in the United States, GroPep Bioreagents (Growpep), Ehime University (Ehime), Walter and Eliza Hall Institute of Medical Research (WEHI). Additional details about panel proteins used in multiplex assays are provided in **Table 1**.

**Table 1.** P. falciparum antigen panel used in multiplex bead-array assays. Information on antigens assessed in this study, including full name, abbreviation, derived protein, alternative names, life-cycle stage of expression, merozoite localisation, and recombinant protein production (producer and protein expression system).

| Antigen |  |  |  | Location & Role |  | Antigen production |  |
| --- | --- | --- | --- | --- | --- | --- | --- |
| Full name | Abbreviation | Derived protein | Other names | Lifecycle stage | Merozoite location | Produced by | Expression system |
| MSP1-19 C-terminal | MSP1<sub>19</sub> | MSP1 |  | Blood | Surface | Burnet | *E. coli* |
| Merozoite surface protein 2 (3D7 allele) | MSP2 | MSP2 |  | Blood | Surface | Growpep | *E.coli* |
| Merozoite surface protein 3 | MSP3 | MSP3 |  | Blood | Surface | Burnet | *E. coli* |
| Merozoite surface protein duffy binding-like 1 | MSPDBL1 | MSPDBL1 | MSP3.4 | Blood | Surface | Ehime | WGCF |
| Merozoite surface protein duffy binding-like 2 | MSPDBL2 | MSPDBL2 | MSP3.8 | Blood | Surface | Ehime | WGCF |
| Merozoite surface protein 4 | MSP4 | MSP4 |  | Blood | Surface | Burnet | HEK 293 |
| Merozoite surface protein 5 | MSP5 | MSP5 |  | Blood | Surface | Burnet | HEK 293 |
| Merozoite surface protein 6 | MSP6 | MSP6 |  | Blood | Surface | Burnet | HEK 293 |
| Merozoite surface protein 7 | MSP7 | MSP7 |  | Blood | Surface | Burnet | HEK 293 |
| Merozoite surface protein 9 | MSP9 | MSP9 | ABRA | Blood | Surface | Ehime | WGCF |
| Merozoite surface protein 10 | MSP10 | MSP10 |  | Blood | Surface | Ehime | WGCF |
| Merozoite surface protein 11 | MSP11 | MSP11 |  | Blood | Organelle (rhoptry) | Ehime | WGCF |
| Glutamate-rich protein | GLURP | GLURP |  | Blood / Pre-erythrocytic | Surface | Genscript | peptide |
| P12 | Pf12 | P12 |  | Multi-stage | Surface | Ehime | WGCF |
| P41 | P41 | P41 |  | Multi-stage | Surface | Ehime | WGCF |
| Glycosylphosphatidylinositol (GPI)-anchored micronemal antigen | GAMA | GAMA |  | Blood | Organelle (microneme) | Ehime | WGCF |
| Apical membrane antigen 1 | AMA1 | AMA1 |  | Blood / Pre-erythrocytic | Organelle (microneme) | Burnet | HEK 293 |
| Erythrocyte binding antigen (140kDa) | EBA140 (RIII-V) | EB140 |  | Blood | Organelle (microneme) | Burnet | *E. coli* |
| Erythrocyte binding antigen (140kDa) | EBA140 (RII) | EBA140 | EBP2 (RII), EBP2,BAEBL | Blood | Organelle (microneme) | NIH | *P. pastoris* |
| Erythrocyte binding antigen (175kDa) | EBA175 (RIII-V) | EBA175 |  | Blood | Organelle (microneme) | Burnet | *E. coli* |
| Erythrocyte binding antigen (175kDa) | EBA175 (RII) | EBA175 |  | Blood | Organelle (microneme) | NIH | *P. pastoris* |
| Erythrocyte binding antigen (181kDa) | EBA181 (RIII-V) | EBA181 | JESEBL | Blood | Organelle (microneme) | Burnet | *E. coli* |
| Cysteine-rich protective antigen | CyRPA | CyRPA |  | Blood | Organelle (microneme) | Ehime | WGCF |
| Reticulocyte binding protein homologue 5-interacting protein | RIPR | RIPR |  | Blood | Organelle (microneme) | Ehime | WGCF |
| Reticulocyte binding protein homologue 2A | Rh2 | Rh2 |  | Blood | Organelle (rhoptry) | Burnet | *E. coli* |
| Reticulocyte Binding Protein-Like Homologue 4 | Rh4 | Rh4 |  | Blood | Organelle (rhoptry) | Burnet | *E. coli* |
| Reticulocyte binding homologue 5 | Rh5 | Rh5 |  | Blood | Organelle (rhoptry) | Ehime | WGCF |
| Rhoptry-associated membrane antigen | RAMA | RAMA |  | Blood | Organelle (rhoptry) | Ehime | WGCF |
| Rhoptry-associated, leucine zipper-like protein 1 | RALP1 | RALP1 |  | Blood | Organelle (rhoptry) | Ehime | WGCF |
| Pfs230 | Pfs230 (D1M) | Pfs230 |  | Transmission | Non-merozoite (gametocyte surface) | Burnet | HEK 293 |
| Circumsporozoite protein | CSP | CSP | CS | Pre-erythrocytic | Non-merozoite (sporozoite surface) | Burnet | HEK 293 |

<sup>1031</sup> **Table 1. *P. falciparum* antigen panel used in multiplex bead-array assays.** Information on antigens assessed in this study, including full name, abbreviation, derived protein, alternative names, life-cycle stage of expression, merozoite localisation, and recombinant protein production (producer and protein expression system).

### *P. falciparum* parasite culturing and merozoite isolation

*P. falciparum* (3D7) parasites were cultured according to established protocols, as described elsewhere [101, 102], where cryo-preserved ring-stage parasites were thawed and added to RBCs at 4% (v/v) haematocrit in media composed of RPMI 1640 (Gibco), HEPES, 0.25% (w/v) Albumax II (GIBCO), 0.5% (v/v) hypoxanthine, 0.18% (w/v) sodium bicarbonate (NaHCO□), and 20 µg/mL gentamicin. Parasite cultures were maintained at 37 °C in a gas mixture of 1% O□, 5% CO□, and 95% N□. Culture media was replaced daily and parasitaemia was monitored by examining Giemsa-stained blood smears.

To isolate merozoites, parasites were synchronised with 5% sorbitol, grown to 10% parasitaemia, then purified for trophozoites by MACS separation (Miltenyl Biotec) and incubated overnight, as described in [12]. Protease inhibitor E64 (10 µg mL) was added to prevent schizont rupture and parasite-enriched cultures were passed through a 1.2 µm filter to collect merozoites, which were quantified using flow cytometry. Wild-type P. falciparum (3D7) parasites were selected as they closely resemble the *P. falciparum* sub-strain (3D7-MBE-008) used to inoculate CHMI participants [103].

### Whole merozoite ELISA

Levels of IgG and IgM to merozoites were quantified using ELISA [13]. Briefly, flat-bottomed 96-well ELISA plates (NUNC MaxiSorp, Thermo Scientific) were coated with freshly isolated P. falciparum merozoites (250,000 parasites per well) suspended in 1 × phosphate-buffered saline (PBS). Plates were incubated overnight at 4 °C. Plates were washed four times with PBS-Tween 0.05% (v/v), which was repeated between each subsequent incubation step. Plates were blocked with 10% (w/v) skim milk in PBS (200 µL per well) and incubated for 2 hours at room temperature (RT). Plasma from CHMI participants was added (1/50 dilution for IgG; 1/2000 for IgM) in 5% (w/v) skim milk-PBS and incubated for 2 hours at RT. To detect antibody isotypes, secondary antibodies were added 1/1000 dilution in 5% (w/v) skim milk-PBS and incubated for 1 hour at RT, with HRP-conjugated goat anti-human IgG (Thermo Scientific) for IgG and HRP-conjugated goat anti-human (Millipore) for IgM. TMB substrate was added and left to develop in the dark at RT. Plates were quenched with 1M H_2_SO_4_ and optical density (OD) absorbance read at 450 nm using a multi-scan GO microplate reader (Thermo Fisher). See Table 2 for details of biological resources.

**Table 2.**
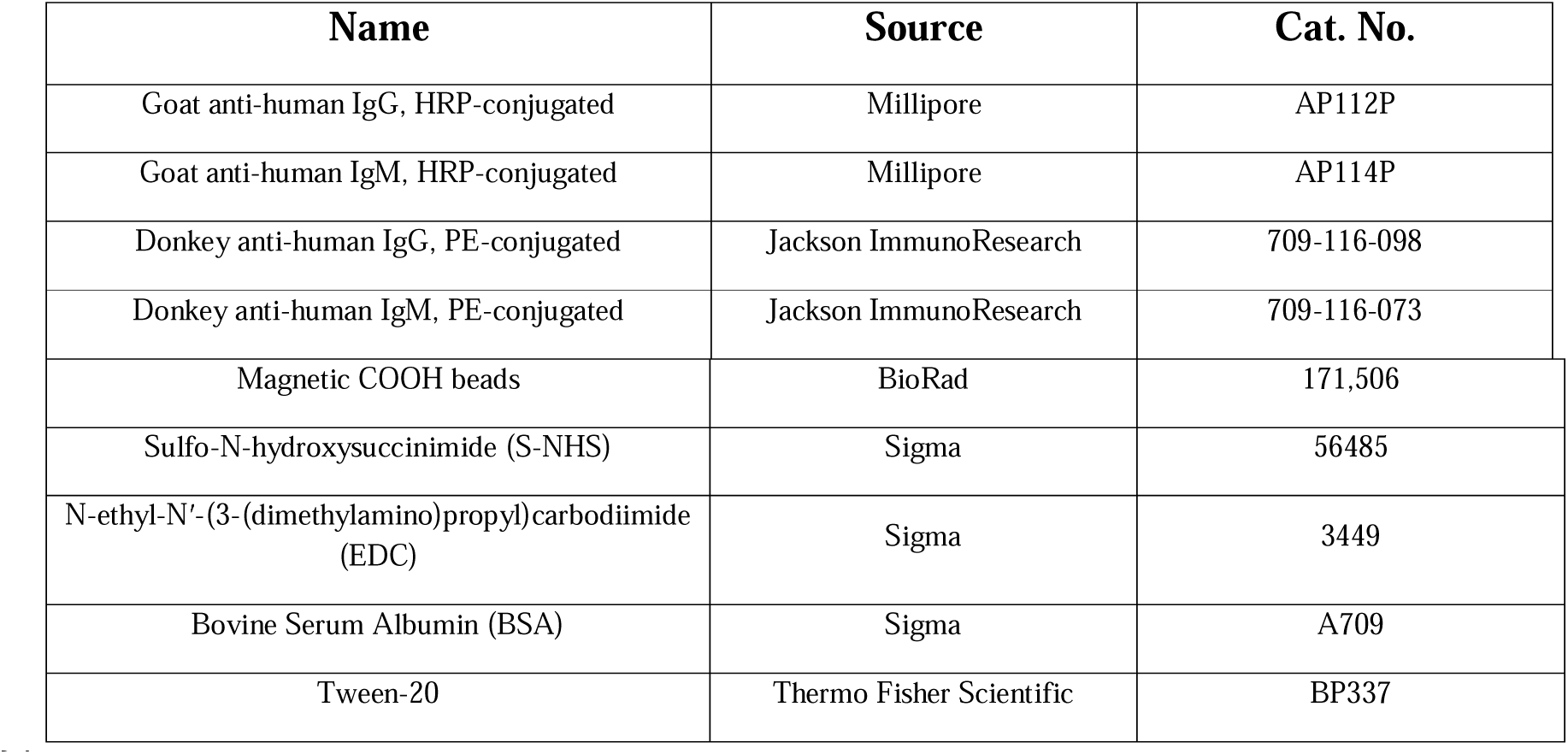
Biological resources used in assays.

Background reactivity (mean OD of blank wells) was subtracted from raw sample OD. Inter-plate variation accounted for by standardising sample OD levels using a plate-specific normalising factor (mean OD plate / mean OD of all plate-specific ODs), found using reactivity from sera from malaria-exposed adults living in Papua New Guinea, which were tested on each plate. Plasma from malaria-naive Melbourne adults was used as negative controls.

### Multiplex antibody assays

Multiplex bead-array assays used *P. falciparum* proteins coupled to magnetic carboxylated microspheres (Luminex) at optimised concentrations, following a standard protocol [104, 105]. To quantify IgG and IgM, assays were performed following a modified version of a protocol previously described [106]. Briefly, protein-coupled beads were incubated with plasma samples in a light-resistant 96-well plate (Greiner Bio-One) for 30 minutes at RT. Beads coupled to each panel protein were added at 0.1 µL per well, diluted in 1 × PBS, 0.1% (w/v) BSA, and Tween-20 (PBT) to make up 50 µL/well. Plasma samples were added at 1:100 dilution in PBT. Plates were washed three times in PBT, which was repeated between each of the subsequent incubations. Secondary antibodies were added to plates at 1:100 dilution in PBT to detect IgG (PE-conjugated donkey anti-human IgG) or IgM (PE-conjugated donkey anti-human IgM), then incubated for 15 minutes at RT. PBT was then added to plates (120 µL/well), which were then incubated for 20 minutes at RT and processed using a BioPlex 200 Luminex System (171000201). Antibody levels were recorded as mean fluorescence intensity (MFI).

Antigen-specific background reactivity (mean of PBT blank wells) was removed from test samples. To account for plate-to-plate variation, each plate included a 10-fold serial titration of WHO P. falciparum reference sera (Prod.No. #10/198), whose reactivity was used to standardise MFI levels across plates. Plasma from malaria-naïve Melbourne (Australia) adults (n=28) were used as additional negative controls.

### Data processing and statistics

All data processing and statistical analyses were performed with R (4.5.2) in RStudio ( 2026.7.0.139). Multiple packages were used to clean and wrangle data (tidyverse, readxl, janitor, scales), perform statistical analysis (ggpubr, rstatix), and visualise trends (ggplot2, ComplexHeatmap). Statistical comparison of antibody levels between two time-points were performed using a paired Wilcoxon signed-rank test, while three or more groups were compared using a Kruskal-Wallis test paired with Dunn’s post-hoc test. Correlations between continuous variables were assessed using Spearman’s rank correlation, with absolute correlation coefficient (rho) values considered as either weak (|rho| <0.5), moderate (0.5 ≤ |rho| < 0.7), or strong (|rho| ≥ 0.7). Clustering of participants by antibody profile was performed using k-means clustering set for three groups (k=3). Statistical significance was defined as p < 0.05 and reported as *p<0.05; **p<0.01; ***p<0.001; ****p<0.0001. Difference in antibody profile of IgG and IgM was assessed using Wilcoxon’s signed rank sum test.

Details of specific tests are provided in each figure legend.

## Declarations

### Ethics approval and consent to participate

Ethics approval for the collection and use of human samples was obtained from the Human Research and Ethics Committee at QIMR-Berghofer Medical Research Institute (P1479, P3444, P3445) and the Alfred Human Research and Ethics Committee at the Burnet Institute (#225/19). All participants provided written informed consent prior to inclusion.

### Consent for publication

Not applicable.

### Availability of data and materials

Data generated for this study and processing scripts are available from the corresponding author upon request, with approval from ethics and regulatory committees where relevant. Materials and reagents used in assays can be provided, depending on availability.

### Competing interests

The authors declare that this research was conducted without conflicts of interest, including any commercial or financial relationships.

### Funding

This work was supported by the National Health and Medical Research Council (NHMRC) of Australia (Investigator Grant and Malaria Synergy grant to JGB), and Australian Centre of Research Excellence in Malaria Elimination (ACREME) Seed Grant to L Reiling; the CSL Centenary Fellowship and the Snow Medical Foundation Fellowship 2022/SF167 to MJ Boyle; NHMRC Investigator Grant to BEB (2016792); Australian Government Research Training Program Scholarship to JW. Burnet Institute is supported by the NHMRC Independent Research Institutes Infrastructure Support Scheme and the Victorian State Government Operational Infrastructure Support. The CHMI trials were previously funded by Medicines for Malaria Venture and Gates Foundation (INV-001965). The conclusions and opinions expressed in this work are those of the author(s) and do not necessarily reflect positions or policies of Gates Foundation, Medicines for Malaria Venture, or other funders. Funders did not play a role in data collection, data analysis, decision to publish, or manuscript preparation.

### Authors’ contributions

JW led writing, performed antibody assays, parasite culturing, data analysis, and visualisation. JGB, LR, JW, HO, MJB, BB conceptualised the study. JW, JGB, HO led data interpretation, with input from LR, LMY, MJB. LR, HO, and JGB raised funding. MJB, BB and CRE provided clinical samples used in this study. BB, JSM conducted the clinical CHMI studies. ET, AT, JGB, LR, KP, HN provided key reagents. All authors contributed to manuscript review.

## Acknowledgements

We thank all participants from the controlled human malaria infection trials evaluated in this study. We also thank all clinicians and support staff at QPharm and University of Sunshine Coast who helped conduct these trials. Red blood cells and human sera were provided by the Australian Red Cross Blood Bank. This study was conducted at the Burnet Institute and QIMR-Berghofer, and we acknowledge the traditional custodians of the lands on which these institutes are located: the Boonwurrung and Wurundjeri Woi-wurrong peoples of the Kulin Nation, and the Turrbal and Jagera peoples.

## Supplementary figures

**Supplementary Figure 1. Whole merozoite antibody seropositivity thresholds.** (A) IgG (top) and IgM (bottom) levels to the whole merozoite surface for malaria-naive Melbourne adults (n =22), CHMI participants (n = 81) at pre-infection, two weeks post-infection, and EOS, and malaria-exposed residents of Papua New Guinea (n = 5). Seropositivity cutoffs (dashed lines) defined as the mean plus two standard deviations above for reactivity of pre-infection CHMI participants (gold) or malaria-naive Melbourne adults (blue). (B) Seroprevalence of IgG (red) and IgM (blue) to whole merozoites at two weeks post-infection and EOS. Seropositivity cutoffs were defined as the mean plus two standard deviations above for reactivity of pre-infection CHMI participants (left) or malaria-naïve Melbourne adults (right).

**Supplementary Figure 2. Merozoite IgG and IgM breadth across participant clusters.** (A) Seroprevalence of IgG (top) and IgM (bottom) to whole merozoite between participant clusters. (B) Seroprevalence of IgG (top) and IgM (bottom) to P. falciparum antigens with antigens ordered according to IgG seroprevalence for cluster 2.

**Supplementary Figure 3. Age associated with more robust merozoite antibody responses following first CHMI.** (A) Age (in years) compared between participant clusters. (B-C) Participants were split into even groups according to age tertile (n=27), including youngest (18-22 years), middle (23-29 years), and oldest (30-55 years). Age groups were compared using IgG and IgM levels to (B) whole merozoites and (C) P. falciparum antigens. (D) Proportion of participants male (brown) or female (fuscia) across participant clusters. (E) Male and female participants were compared using IgG and IgM levels to whole merozoites at two weeks post-infection and EOS. Group comparisons in A, B, and C performed using Kruskal-Wallis test paired with a Dunn test, whereas comparisons in E were performed using a Mann-Whitney U test. Only significant (p < 0.05) comparisons shown.

## Notes

### Competing Interest Statement

The authors have declared no competing interest.

## References

1. World Health Organization. World malaria report 2025: Addressing the threat of antimalarial drug resistance. Global report. World Health Organization; 2025.

2. Marsh K, Kinyanjui S. Immune effector mechanisms in malaria. Parasite Immunol. 2006;28:51–60. 10.1111/j.1365-3024.2006.00808.x.

3. Doolan DL, Dobaño C, Baird JK. Acquired Immunity to Malaria. Clin Microbiol Rev. 2009;22:13–36. 10.1128/CMR.00025-08.

4. Gupta S, Snow RW, Donnelly CA, Marsh K, Newbold C. Immunity to non-cerebral severe malaria is acquired after one or two infections. Nat Med. 1999;5:340–3. 10.1038/6560.

5. Fowkes FJI, Richards JS, Simpson JA, Beeson JG. The Relationship between Anti-merozoite Antibodies and Incidence of Plasmodium falciparum Malaria: A Systematic Review and Meta-analysis. PLOS Med. 2010;7:e1000218. 10.1371/journal.pmed.1000218.

6. Richards JS, Stanisic DI, Fowkes FJI, Tavul L, Dabod E, Thompson JK, et al. Association between Naturally Acquired Antibodies to Erythrocyte-Binding Antigens of Plasmodium falciparum and Protection from Malaria and High-Density Parasitemia. Clin Infect Dis. 2010;51:e50–60. 10.1086/656413.

7. Richards JS, Arumugam TU, Reiling L, Healer J, Hodder AN, Fowkes FJI, et al. Identification and Prioritization of Merozoite Antigens as Targets of Protective Human Immunity to Plasmodium falciparum Malaria for Vaccine and Biomarker Development. J Immunol. 2013;191:795–809. 10.4049/jimmunol.1300778.

8. Reiling L, Boyle MJ, White MT, Wilson DW, Feng G, Weaver R, et al. Targets of complement-fixing antibodies in protective immunity against malaria in children. Nat Commun. 2019;10:610. 10.1038/s41467-019-08528-z.

9. Hill DL, Wilson DW, Sampaio NG, Eriksson EM, Ryg-Cornejo V, Harrison GLA, et al. Merozoite Antigens of Plasmodium falciparum Elicit Strain-Transcending Opsonizing Immunity. Infect Immun. 2016;84:2175–84. 10.1128/IAI.00145-16.

10. Stanisic DI, Richards JS, McCallum FJ, Michon P, King CL, Schoepflin S, et al. Immunoglobulin G Subclass-Specific Responses against Plasmodium falciparum Merozoite Antigens Are Associated with Control of Parasitemia and Protection from Symptomatic Illness. Infect Immun. 2009;77:1165–74. 10.1128/iai.01129-08.

11. Osier FH, Feng G, Boyle MJ, Langer C, Zhou J, Richards JS, et al. Opsonic phagocytosis of Plasmodium falciparum merozoites: mechanism in human immunity and a correlate of protection against malaria. BMC Med. 2014;12:108. 10.1186/1741-7015-12-108.

12. Boyle MJ, Reiling L, Feng G, Langer C, Osier FH, Aspeling-Jones H, et al. Human Antibodies Fix Complement to Inhibit Plasmodium falciparum Invasion of Erythrocytes and Are Associated with Protection against Malaria. Immunity. 2015;42:580–90. 10.1016/j.immuni.2015.02.012.

13. Boyle MJ, Chan JA, Handayuni I, Reiling L, Feng G, Hilton A, et al. IgM in human immunity to Plasmodium falciparum malaria. Sci Adv. 2019;5:eaax4489. 10.1126/sciadv.aax4489.

14. Beeson JG, Kurtovic L, Valim C, Asante KP, Boyle MJ, Mathanga D, et al. The RTS,S malaria vaccine: Current impact and foundation for the future. Sci Transl Med. 2022;14:eabo6646. 10.1126/scitranslmed.abo6646.

15. Hassert M, Harty JT. The endemic dilemma: why whole sporozoite malaria vaccine efficacy falls in the field. Trends Parasitol. 2026;42:105–14. 10.1016/j.pt.2025.12.003.

16. Beeson JG, Kurtovic L, Dobaño C, Opi DH, Chan J-A, Feng G, et al. Challenges and strategies for developing efficacious and long-lasting malaria vaccines. Sci Transl Med. 2019;11:eaau1458. 10.1126/scitranslmed.aau1458.

17. Osier FHA, Fegan G, Polley SD, Murungi L, Verra F, Tetteh KKA, et al. Breadth and Magnitude of Antibody Responses to Multiple Plasmodium falciparum Merozoite Antigens Are Associated with Protection from Clinical Malaria. Infect Immun. 2008;76:2240–8. 10.1128/IAI.01585-07.

18. Roussilhon C, Oeuvray C, Müller-Graf C, Tall A, Rogier C, Trape J-F, et al. Long-Term Clinical Protection from Falciparum Malaria Is Strongly Associated with IgG3 Antibodies to Merozoite Surface Protein 3. PLoS Med. 2007;4:e320. 10.1371/journal.pmed.0040320.

19. Afolabi BM, Salako LA, Mafe AG, Ovwigho UB, Rabiu KA, Sanyaolu NO, et al. Malaria in the first 6 months of life in urban African infants with anemia. Am J Trop Med Hyg. 2001;65:822–7. 10.4269/ajtmh.2001.65.822.

20. Snow RW, Marsh K. New insights into the epidemiology of malaria relevant for disease control. Br Med Bull. 1998;54:293–309. 10.1093/oxfordjournals.bmb.a011689.

21. McGuinness D, Koram K, Bennett S, Wagner G, Nkrumah F, Riley E. Clinical case definitions for malaria: clinical malaria associated with very low parasite densities in African infants. Trans R Soc Trop Med Hyg. 1998;92:527–31. 10.1016/S0035-9203(98)90902-6.

22. Surendra H, Supargiyono, Ahmad RA, Kusumasari RA, Rahayujati TB, Damayanti SY, et al. Using health facility-based serological surveillance to predict receptive areas at risk of malaria outbreaks in elimination areas. BMC Med. 2020;18:9. 10.1186/s12916-019-1482-7.

23. Wu L, Mwesigwa J, Affara M, Bah M, Correa S, Hall T, et al. Antibody responses to a suite of novel serological markers for malaria surveillance demonstrate strong correlation with clinical and parasitological infection across seasons and transmission settings in The Gambia. BMC Med. 2020;18:304. 10.1186/s12916-020-01724-5.

24. Lim PS, Bernal J, Karunarathne J, Longley RJ. Malaria serology for surveillance, insights into immunity, and vaccine development. Infect Immun. 2026;94:e00031–26. 10.1128/iai.00031-26.

25. Sauerwein RW, Roestenberg M, Moorthy VS. Experimental human challenge infections can accelerate clinical malaria vaccine development. Nat Rev Immunol. 2011;11:57–64. 10.1038/nri2902.

26. Yman V, White MT, Asghar M, Sundling C, Sondén K, Draper SJ, et al. Antibody responses to merozoite antigens after natural Plasmodium falciparum infection: kinetics and longevity in absence of re-exposure. BMC Med. 2019;17:22. 10.1186/s12916-019-1255-3.

27. White MT, Griffin JT, Akpogheneta O, Conway DJ, Koram KA, Riley EM, et al. Dynamics of the Antibody Response to Plasmodium falciparum Infection in African Children. J Infect Dis. 2014;210:1115–22. 10.1093/infdis/jiu219.

28. Tijani MK, Babalola OA, Odaibo AB, Anumudu CI, Asinobi AO, Morenikeji OA, et al. Acquisition, maintenance and adaptation of invasion inhibitory antibodies against Plasmodium falciparum invasion ligands involved in immune evasion. PLoS One. 2017;12:e0182187. 10.1371/journal.pone.0182187.

29. van den Hoogen LL, Walk J, Oulton T, Reuling IJ, Reiling L, Beeson JG, et al. Antibody Responses to Antigenic Targets of Recent Exposure Are Associated With Low-Density Parasitemia in Controlled Human Plasmodium falciparum Infections. Front Microbiol. 2019;9:3300. 10.3389/fmicb.2018.03300.

30. Stanisic DI, McCarthy JS, Good MF. Controlled Human Malaria Infection: Applications, Advances, and Challenges. Infect Immun. 2018;86:e00479–17. 10.1128/iai.00479-17.

31. Scholzen A, Sauerwein RW. Immune activation and induction of memory: lessons learned from controlled human malaria infection with Plasmodium falciparum. Parasitology. 2016;143:224–35. 10.1017/S0031182015000761.

32. Biswas S, Choudhary P, Elias SC, Miura K, Milne KH, de Cassan SC, et al. Assessment of humoral immune responses to blood-stage malaria antigens following ChAd63-MVA immunization, controlled human malaria infection and natural exposure. PLoS One. 2014;9:e107903. 10.1371/journal.pone.0107903.

33. Hall CE, Hagan LM, Bergmann-Leitner E, Tosh DM, Bennett JW, Regules JA, et al. Mosquito Bite-Induced Controlled Human Malaria Infection with Plasmodium vivax or P. falciparum Generates Immune Responses to Homologous and Heterologous Preerythrocytic and Erythrocytic Antigens. Infect Immun. 2019;87:e00541–18. 10.1128/IAI.00541-18.

34. Sedegah M, Hollingdale MR, Farooq F, Ganeshan H, Belmonte M, Huang J, et al. Controlled Human Malaria Infection (CHMI) differentially affects cell-mediated and antibody responses to CSP and AMA1 induced by adenovirus vaccines with and without DNA-priming. Hum Vaccin Immunother. 2015;11:2705–15. 10.1080/21645515.2015.1019186.

35. Payne RO, Milne KH, Elias SC, Edwards NJ, Douglas AD, Brown RE, et al. Demonstration of the Blood-Stage Plasmodium falciparum Controlled Human Malaria Infection Model to Assess Efficacy of the P. falciparum Apical Membrane Antigen 1 Vaccine, FMP2.1/AS01. J Infect Dis. 2016;213:1743–51. 10.1093/infdis/jiw039.

36. Salkeld J, Themistocleous Y, Barrett JR, Mitton CH, Rawlinson TA, Payne RO, et al. Repeat controlled human malaria infection of healthy UK adults with blood-stage Plasmodium falciparum: Safety and parasite growth dynamics. Front Immunol. 2022;13:984323. 10.3389/fimmu.2022.984323.

37. Hodgson SH, Llewellyn D, Silk SE, Milne KH, Elias SC, Miura K, et al. Changes in Serological Immunology Measures in UK and Kenyan Adults Post-controlled Human Malaria Infection. Front Microbiol. 2016;7:1604. 10.3389/fmicb.2016.01604.

38. Sheehy SH, Duncan CJ, Elias SC, Choudhary P, Biswas S, Halstead FD, et al. ChAd63-MVA–vectored Blood-stage Malaria Vaccines Targeting MSP1 and AMA1: Assessment of Efficacy Against Mosquito Bite Challenge in Humans. Mol Ther. 2012;20:2355–68. 10.1038/mt.2012.223.

39. Elias SC, Choudhary P, de Cassan SC, Biswas S, Collins KA, Halstead FD, et al. Analysis of human B-cell responses following ChAd63-MVA MSP1 and AMA1 immunization and controlled malaria infection. Immunology. 2014;141:628–44. 10.1111/imm.12226.

40. Nkumama IN, Ogwang R, Odera D, Musasia F, Mwai K, Nyamako L, et al. Breadth of Fc-mediated effector function correlates with clinical immunity following human malaria challenge. Immunity. 2024;57:1215–1224.e6. 10.1016/j.immuni.2024.05.001.

41. Obiero JM, Shekalaghe S, Hermsen CC, Mpina M, Bijker EM, Roestenberg M, et al. Impact of Malaria Preexposure on Antiparasite Cellular and Humoral Immune Responses after Controlled Human Malaria Infection. Infect Immun. 2015;83:2185–96. 10.1128/IAI.03069-14.

42. Hickey BW, Lumsden JM, Reyes S, Sedegah M, Hollingdale MR, Freilich DA, et al. Mosquito bite immunization with radiation-attenuated Plasmodium falciparum sporozoites: safety, tolerability, protective efficacy and humoral immunogenicity. Malar J. 2016;15:377. 10.1186/s12936-016-1435-y.

43. Turner L, Wang CW, Lavstsen T, Mwakalinga SB, Sauerwein RW, Hermsen CC, et al. Antibodies against PfEMP1, RIFIN, MSP3 and GLURP Are Acquired during Controlled Plasmodium falciparum Malaria Infections in Naïve Volunteers. PLoS One. 2011;6:e29025. 10.1371/journal.pone.0029025.

44. Chan J-A, Loughland JR, de Labastida Rivera F, SheelaNair A, Andrew DW, Dooley NL, et al. Th2-like T Follicular Helper Cells Promote Functional Antibody Production during Plasmodium falciparum Infection. Cell Rep Med. 2020;1:100157. 10.1016/j.xcrm.2020.100157.

45. Minassian AM, Silk SE, Barrett JR, Nielsen CM, Miura K, Diouf A, et al. Reduced blood-stage malaria growth and immune correlates in humans following RH5 vaccination. Med. 2021;2:701–719.e19. 10.1016/j.medj.2021.03.014.

46. Gómez-Pérez GP, Vidal M, Campo JJ, Moncunill G, Jimenez A, Vázquez-Santiago M, et al. IgM, IgG, and IgG Subclass Antibody Responses to Plasmodium falciparum Proteins in Naïve, Malaria-Vaccinated and Semi-Immune Volunteers after Controlled Human Malaria Infection. Am J Trop Med Hyg. 2025;113:1235–48. 10.4269/ajtmh.25-0384.

47. Beeson JG, Drew DR, Boyle MJ, Feng G, Fowkes FJI, Richards JS. Merozoite surface proteins in red blood cell invasion, immunity and vaccines against malaria. FEMS Microbiol Rev. 2016;40:343–72. 10.1093/femsre/fuw001.

48. Osier FH, Mackinnon MJ, Crosnier C, Fegan G, Kamuyu G, Wanaguru M, et al. New antigens for a multicomponent blood-stage malaria vaccine. Sci Transl Med. 2014;6:247ra102. 10.1126/scitranslmed.3008705.

49. Reiling L, Richards JS, Fowkes FJI, Barry AE, Triglia T, Chokejindachai W, et al. Evidence That the Erythrocyte Invasion Ligand PfRh2 is a Target of Protective Immunity against Plasmodium falciparum Malaria. J Immunol. 2010;185:6157–67. 10.4049/jimmunol.1001555.

50. Collins KA, Wang CYT, Adams M, Mitchell H, Rampton M, Elliott S, et al. A controlled human malaria infection model enabling evaluation of transmission-blocking interventions. J Clin Invest. 2018;128:1551–62. 10.1172/JCI98012.

51. Bousema T, Roeffen W, Meijerink H, Mwerinde H, Mwakalinga S, van Gemert G-J, et al. The Dynamics of Naturally Acquired Immune Responses to Plasmodium falciparum Sexual Stage Antigens Pfs230 & Pfs48/45 in a Low Endemic Area in Tanzania. PLoS One. 2010;5:e14114. 10.1371/journal.pone.0014114.

52. Skinner J, Huang C-Y, Waisberg M, Felgner PL, Doumbo OK, Ongoiba A, et al. Plasmodium falciparum Gametocyte-Specific Antibody Profiling Reveals Boosting through Natural Infection and Identifies Potential Markers of Gametocyte Exposure. Infect Immun. 2015;83:4229–36. 10.1128/IAI.00644-15.

53. Macià D, Campo JJ, Moncunill G, Jairoce C, Nhabomba AJ, Mpina M, et al. Strong off-target antibody reactivity to malarial antigens induced by RTS,S/AS01E vaccination is associated with protection. JCI Insight. 2022;7:e158030. 10.1172/jci.insight.158030.

54. Ferrer P, Berry AA, Bucsan AN, Prajapati SK, Krishnan K, Barbeau MC, et al. Repeat controlled human Plasmodium falciparum infections delay bloodstream patency and reduce symptoms. Nat Commun. 2024;15:5194. 10.1038/s41467-024-49041-2.

55. Gilson PR, Nebl T, Vukcevic D, Moritz RL, Sargeant T, Speed TP, et al. Identification and Stoichiometry of Glycosylphosphatidylinositol-anchored Membrane Proteins of the Human Malaria Parasite Plasmodium falciparum. Mol Cell Proteomics. 2006;5:1286–99. 10.1074/mcp.M600035-MCP200.

56. Yagi M, Palacpac NMQ, Ito K, Oishi Y, Itagaki S, Balikagala B, et al. Antibody titres and boosting after natural malaria infection in BK-SE36 vaccine responders during a follow-up study in Uganda. Sci Rep. 2016;6:34363. 10.1038/srep34363.

57. Aide P, Dobaño C, Sacarlal J, Aponte JJ, Mandomando I, Guinovart C, et al. Four year immunogenicity of the RTS,S/AS02A malaria vaccine in Mozambican children during a phase IIb trial. Vaccine. 2011;29:6059–67. 10.1016/j.vaccine.2011.03.041.

58. Pombo DJ, Lawrence G, Hirunpetcharat C, Rzepczyk C, Bryden M, Cloonan N, et al. Immunity to malaria after administration of ultra-low doses of red cells infected with Plasmodium falciparum. Lancet. 2002;360:610–7. 10.1016/S0140-6736(02)09784-2.

59. Molineaux L, Träuble M, Collins WE, Jeffery GM, Dietz K. Malaria therapy reinoculation data suggest individual variation of an innate immune response and independent acquisition of antiparasitic and antitoxic immunities. Trans R Soc Trop Med Hyg. 2002;96:205–9. 10.1016/S0035-9203(02)90308-1.

60. Arévalo-Herrera M, Forero-Peña DA, Rubiano K, Gómez-Hincapie J, Martínez NL, Lopez-Perez M, et al. Plasmodium vivax Sporozoite Challenge in Malaria-Naïve and Semi-Immune Colombian Volunteers. PLoS One. 2014;9:e99754. 10.1371/journal.pone.0099754.

61. Arumugam TU, Takeo S, Yamasaki T, Thonkukiatkul A, Miura K, Otsuki H, et al. Discovery of GAMA, a Plasmodium falciparum Merozoite Micronemal Protein, as a Novel Blood-Stage Vaccine Candidate Antigen. Infect Immun. 2011;79:4523–32. 10.1128/IAI.05412-11.

62. Hinds L, Green JL, Knuepfer E, Grainger M, Holder AA. Novel Putative Glycosylphosphatidylinositol-Anchored Micronemal Antigen of Plasmodium falciparum That Binds to Erythrocytes. Eukaryot Cell. 2009;8:1869–79. 10.1128/EC.00218-09.

63. Nagaoka H, Kanoi BN, Jinoka K, Morita M, Arumugam TU, Palacpac NMQ, et al. The N-Terminal Region of Plasmodium falciparum MSP10 Is a Target of Protective Antibodies in Malaria and Is Important for PfGAMA/PfMSP10 Interaction. Front Immunol. 2019;10:2669. 10.3389/fimmu.2019.02669.

64. Maskus DJ, Bethke S, Seidel M, Kapelski S, Addai-Mensah O, Boes A, et al. Isolation, production and characterization of fully human monoclonal antibodies directed to Plasmodium falciparum MSP10. Malar J. 2015;14:276. 10.1186/s12936-015-0797-x.

65. Black CG, Wang L, Wu T, Coppel RL. Apical location of a novel EGF-like domain-containing protein of *Plasmodium falciparum*. Mol Biochem Parasitol. 2003;127:59–68. 10.1016/S0166-6851(02)00308-0.

66. Yman V, Tuju J, White MT, Kamuyu G, Mwai K, Kibinge N, et al. Distinct kinetics of antibodies to 111 Plasmodium falciparum proteins identifies markers of recent malaria exposure. Nat Commun. 2022;13:331. 10.1038/s41467-021-27863-8.

67. Natama HM, Salkeld J, Somé A, Soremekun S, Diallo S, Traoré O, et al. Safety and efficacy of the blood-stage malaria vaccine RH5.1/Matrix-M in Burkina Faso: interim results of a double-blind, randomised, controlled, phase 2b trial in children. Lancet Infect Dis. 2025;25:495–506. 10.1016/S1473-3099(24)00752-7.

68. University of Oxford. A Phase Ib Age De-escalation, Open Label Study of the Safety and Immunogenicity of the Multi-stage Malaria Vaccine Candidate R21 + RH5.1 + R78C in Matrix-M^TM^ in Adults Aged 18-35 Years and Children Aged 5-17 Months in Burkina Faso. Clinical trial registration. clinicaltrials.gov; 2026.

69. Wang LT, Cooper AJR, Farrell B, Miura K, Diouf A, Müller-Sienerth N, et al. Natural malaria infection elicits rare but potent neutralizing antibodies to the blood-stage antigen RH5. Cell. 2024;187:4981–4995.e14. 10.1016/j.cell.2024.06.037.

70. Douglas AD, Williams AR, Illingworth JJ, Kamuyu G, Biswas S, Goodman AL, et al. The blood-stage malaria antigen PfRH5 is susceptible to vaccine-inducible cross-strain neutralizing antibody. Nat Commun. 2011;2:601. 10.1038/ncomms1615.

71. Tran TM, Ongoiba A, Coursen J, Crosnier C, Diouf A, Huang C-Y, et al. Naturally Acquired Antibodies Specific for Plasmodium falciparum Reticulocyte-Binding Protein Homologue 5 Inhibit Parasite Growth and Predict Protection From Malaria. J Infect Dis. 2014;209:789–98. 10.1093/infdis/jit553.

72. Seager BA, Lim PS, Xiao X, Lai KH, Feufack-Donfack LB, Dass S, et al. PTRAMP, CSS and Ripr form a conserved complex required for merozoite invasion of Plasmodium species into erythrocytes. Nat Commun. 2026;17:1780. 10.1038/s41467-026-68486-1.

73. Healer J, Wong W, Thompson JK, He W, Birkinshaw RW, Miura K, et al. Neutralising antibodies block the function of Rh5/Ripr/CyRPA complex during invasion of Plasmodium falciparum into human erythrocytes. Cell Microbiol. 2019;21:e13030. 10.1111/cmi.13030.

74. Mian SY, Somanathan A, Chaddha K, Pandey AK, Singh H, Krishna S, et al. Plasmodium falciparum Cysteine-Rich Protective Antigen (CyRPA) Elicits Detectable Levels of Invasion-Inhibitory Antibodies during Natural Infection in Humans. Infect Immun. 2022;90:e00377–21. 10.1128/IAI.00377-21.

75. Partey FD, Castberg FC, Sarbah EW, Silk SE, Awandare GA, Draper SJ, et al. Kinetics of antibody responses to PfRH5-complex antigens in Ghanaian children with Plasmodium falciparum malaria. PLoS One. 2018;13:e0198371. 10.1371/journal.pone.0198371.

76. Hope IA, Hall R, Simmons DL, Hyde JE, Scaife JG. Evidence for immunological cross-reaction between sporozoites and blood stages of a human malaria parasite. Nature. 1984;308:191–4. 10.1038/308191a0.

77. Coppel RL, Favaloro JM, Crewther PE, Burkot TR, Bianco AE, Stahl HD, et al. A blood stage antigen of Plasmodium falciparum shares determinants with the sporozoite coat protein. Proc Natl Acad Sci U A. 1985;82:5121–5. 10.1073/pnas.82.15.5121.

78. Ardeshir F, Howard RF, Viriyakosol S, Arad O, Reese RT. Cross-reactive asparagine-rich determinants shared between several blood-stage antigens of *Plasmodium falciparum* and the circumsporozoite protein. Mol Biochem Parasitol. 1990;40:113–28. 10.1016/0166-6851(90)90085-Z.

79. Hou MM, Harding AC, Barber NM, Kundu P, Bach FA, Salkeld J, et al. Development of clinical immunity to Plasmodium vivax following repeat controlled human malaria infection. Nat Commun. 2025;16:8385. 10.1038/s41467-025-63104-y.

80. Mukhiya R, Fleischmann WA, Loughland JR, Chan J-A, de Labastida Rivera F, Andrew D, et al. Heterogeneity of the human immune response to malaria infection and vaccination driven by latent cytomegalovirus infection. eBioMedicine. 2024;109:105419. 10.1016/j.ebiom.2024.105419.

81. Valletta JJ, Addy JWG, Reid AJ, Ndungu FM, Bediako Y, Mwacharo J, et al. Individual-level variations in malaria susceptibility and acquisition of clinical protection. Wellcome Open Res. 2022;6:22. 10.12688/wellcomeopenres.16524.3.

82. Trape J-F, Diagne N, Diene-Sarr F, Faye J, Dieye-Ba F, Bassène H, et al. One hundred malaria attacks since birth. A longitudinal study of African children and young adults exposed to high malaria transmission. eClinicalMedicine. 2023;67:102379. 10.1016/j.eclinm.2023.102379.

83. Gaur AH, McCarthy JS, Panetta JC, Dallas RH, Woodford J, Tang L, et al. Safety, tolerability, pharmacokinetics, and antimalarial efficacy of a novel Plasmodium falciparum ATP4 inhibitor SJ733: a first-in-human and induced blood-stage malaria phase 1a/b trial. Lancet Infect Dis. 2020;20:964–75. 10.1016/S1473-3099(19)30611-5.

84. Abd-Rahman AN, Kaschek D, Kümmel A, Webster R, Potter AJ, Odedra A, et al. Characterizing the pharmacological interaction of the antimalarial combination artefenomel-piperaquine in healthy volunteers with induced blood-stage Plasmodium falciparum to predict efficacy in patients with malaria. BMC Med. 2024;22:563. 10.1186/s12916-024-03787-0.

85. McCarthy JS, Donini C, Chalon S, Woodford J, Marquart L, Collins KA, et al. A Phase 1, Placebo-controlled, Randomized, Single Ascending Dose Study and a Volunteer Infection Study to Characterize the Safety, Pharmacokinetics, and Antimalarial Activity of the Plasmodium Phosphatidylinositol 4-Kinase Inhibitor MMV390048. Clin Infect Dis. 2020;71:e657–64. 10.1093/cid/ciaa368.

86. Barber BE, Abd-Rahman AN, Webster R, Potter AJ, Llewellyn S, Marquart L, et al. Characterizing the Blood-Stage Antimalarial Activity of Tafenoquine in Healthy Volunteers Experimentally Infected With Plasmodium falciparum. Clin Infect Dis. 2023;76:1919–27. 10.1093/cid/ciad075.

87. Soumare HM, Blanken SL, Ahmad A, Ooko M, Gaye PM, Jadama L, et al. Asymptomatic school children and adults are important for the human infectious reservoir for Plasmodium falciparum malaria in an area of low endemicity in The Gambia. J Infect. 2025;91:106507. 10.1016/j.jinf.2025.106507.

88. Slater HC, Ross A, Felger I, Hofmann NE, Robinson L, Cook J, et al. The temporal dynamics and infectiousness of subpatent Plasmodium falciparum infections in relation to parasite density. Nat Commun. 2019;10:1433. 10.1038/s41467-019-09441-1.

89. Akpogheneta OJ, Duah NO, Tetteh KKA, Dunyo S, Lanar DE, Pinder M, et al. Duration of Naturally Acquired Antibody Responses to Blood-Stage Plasmodium falciparum Is Age Dependent and Antigen Specific. Infect Immun. 2008;76:1748–55. 10.1128/IAI.01333-07.

90. Greenhouse B, Ho B, Hubbard A, Njama-Meya D, Narum DL, Lanar DE, et al. Antibodies to Plasmodium falciparum Antigens Predict a Higher Risk of Malaria But Protection From Symptoms Once Parasitemic. J Infect Dis. 2011;204:19–26. 10.1093/infdis/jir223.

91. Rono J, Osier FHA, Olsson D, Montgomery S, Mhoja L, Rooth I, et al. Breadth of Anti-Merozoite Antibody Responses Is Associated With the Genetic Diversity of Asymptomatic Plasmodium falciparum Infections and Protection Against Clinical Malaria. Clin Infect Dis. 2013;57:1409–16. 10.1093/cid/cit556.

92. Friedman-Klabanoff DJ, Jensen TL, Lyke KE, Laurens MB, Silva JC, Stucke EM, et al. Controlled human malaria infection with NF54 and 7G8 strains elicit differential antibody responses to Plasmodium falciparum peptides. Front Immunol. 2025;16:1641280. 10.3389/fimmu.2025.1641280.

93. Loughland JR, Dooley NL, Pava Z, SheelaNair A, Andrew DW, Tipping P, et al. Age is an intrinsic driver of inflammatory responses to malaria. Nat Commun. 2025;16:8665. 10.1038/s41467-025-63638-1.

94. Rodriguez-Barraquer I, Arinaitwe E, Jagannathan P, Kamya MR, Rosenthal PJ, Rek J, et al. Quantification of anti-parasite and anti-disease immunity to malaria as a function of age and exposure. eLife. 2018;7:e35832. 10.7554/eLife.35832.

95. Duncombe CJ, Hergott DEB, Staubus W, Balke-Buijs M, Kublin JG, Duffy PE, et al. Sex-based differences in Plasmodium infection in the control groups of controlled human malaria infection trials in malaria-naive populations in the USA and the Netherlands: a pooled analysis. Lancet Microbe. 2026;7:101265. 10.1016/j.lanmic.2025.101265.

96. Briggs J, Teyssier N, Nankabirwa JI, Rek J, Jagannathan P, Arinaitwe E, et al. Sex-based differences in clearance of chronic Plasmodium falciparum infection. eLife. 2020;9:e59872. 10.7554/eLife.59872.

97. Kc N, Church LWP, Riyahi P, Chakravarty S, Seder RA, Epstein JE, et al. Increased levels of anti-PfCSP antibodies in post-pubertal females versus males immunized with PfSPZ Vaccine does not translate into increased protective efficacy. Front Immunol. 2022;13:1006716. 10.3389/fimmu.2022.1006716.

98. Kurtovic L, Feng G, Hysa A, Haghiri A, O’Flaherty K, Wines BD, et al. Antibody mechanisms of protection against malaria in RTS,S-vaccinated children: a post-hoc serological analysis of phase 2 trial. Lancet Microbe. 2024;5:100898. 10.1016/S2666-5247(24)00130-7.

99. McCarthy JS, Marjason J, Elliott S, Fahey P, Bang G, Malkin E, et al. A Phase 1 Trial of MSP2-C1, a Blood-Stage Malaria Vaccine Containing 2 Isoforms of MSP2 Formulated with Montanide® ISA 720. PLoS One. 2011;6:e24413. 10.1371/journal.pone.0024413.

100. Rockett RJ, Tozer SJ, Peatey C, Bialasiewicz S, Whiley DM, Nissen MD, et al. A real-time, quantitative PCR method using hydrolysis probes for the monitoring of Plasmodium falciparum load in experimentally infected human volunteers. Malar J. 2011;10:48. 10.1186/1475-2875-10-48.

101. Beeson JG, Brown GV, Molyneux ME, Mhango C, Dzinjalamala F, Rogerson SJ. Plasmodium falciparum Isolates from Infected Pregnant Women and Children Are Associated with Distinct Adhesive and Antigenic Properties. J Infect Dis. 1999;180:464–72. 10.1086/314899.

102. Maier AG, Rug M. In Vitro Culturing Plasmodium falciparum Erythrocytic Stages. In: Menard R, editor. Malaria: Methods and Protocols. 2nd edition. Humana Press; 2013. p. 3–15. 10.1007/978-1-62703-026-7_1.

103. Woolley SD, Fernandez M, Rebelo M, Llewellyn SA, Marquart L, Amante FH, et al. Development and evaluation of a new Plasmodium falciparum 3D7 blood stage malaria cell bank for use in malaria volunteer infection studies. Malar J. 2021;20:93. 10.1186/s12936-021-03627-z.

104. Ubillos I, Jiménez A, Vidal M, Bowyer PW, Gaur D, Dutta S, et al. Optimization of incubation conditions of Plasmodium falciparum antibody multiplex assays to measure IgG, IgG1–4, IgM and IgE using standard and customized reference pools for sero-epidemiological and vaccine studies. Malar J. 2018;17:219. 10.1186/s12936-018-2369-3.

105. Mazhari R, Brewster J, Fong R, Bourke C, Liu ZSJ, Takashima E, et al. A comparison of non-magnetic and magnetic beads for measuring IgG antibodies against Plasmodium vivax antigens in a multiplexed bead-based assay using Luminex technology (Bio-Plex 200 or MAGPIX). PLoS One. 2020;15:e0238010. 10.1371/journal.pone.0238010.

106. Longley RJ, White MT, Takashima E, Brewster J, Morita M, Harbers M, et al. Development and validation of serological markers for detecting recent Plasmodium vivax infection. Nat Med. 2020;26:741–9. 10.1038/s41591-020-0841-4.

