## Supplementary Figures for "Hierarchy of immunogenicity of *Plasmodium falciparum* merozoite antigens and vaccine candidates in first human infection"

|  |  |  |  |  | Sex, n (%) | | Age (y) |
| --- | --- | --- | --- | --- | --- | --- | --- |
| Trial registration | Study | Drug administered  at peak infection | Cohorts (n) | Participants (n) | **Female Male** | | **Median (range)** |
| ACTRN12620000995976 | CTM2002LIH | Tafenoquine | 3 | 18 | 9 (50) | 9 (50) | 31 (19–56) |
| NCT02431637 | QP14C21 | PQP | 2 | 4 | 4 (100) | 0 (0) | 26 (21 - 31) |
| NCT02431650 | QP15C05 | PQP + OZ439 or primaquine | 3 | 6 | 0 (0) | 6 (100) | 24 (20 - 34) |
| NCT02867059 | QP15C20 | SJ733 | 4 | 15 | 0 (0) | 15 (100) | 23 (18 – 52) |
| NCT02783833 | QP16C04 | MMV390048 | 2 | 15 | 0 (0) | 15 (100) | 29 (20 - 48) |
| NCT03542149 | QP17C19 | PQP | 3 | 23 | 6 (26) | 17 (74) | 22 (18 - 53) |
|  |  | **Total** | 17 | 81 | 19 (23) | 62 (77) | 24 (18 – 56) |

**Supplementary Table 1. Characteristics of clinical studies and participants assessed in this study**. Summary of included studies, including trial registration number, study name, antimalarial drug administered at peak infection (7/8 days post-inoculation, number of cohorts and participants, participant sex (n ( %)), and age (years, median (range)). PQP = piperaquine phosphate; OZ439 is also known as artefenomel.

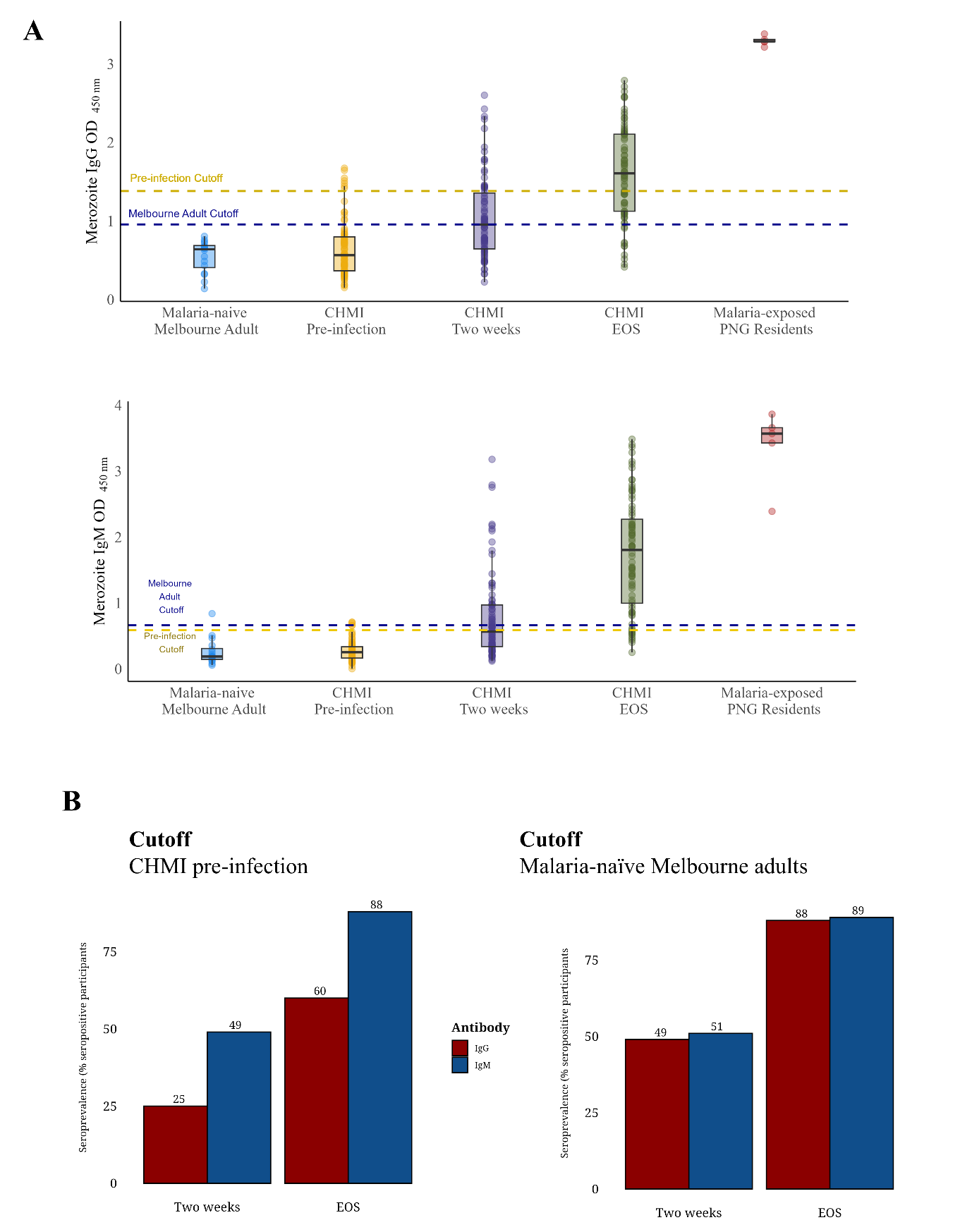

**Supplementary Figure 1. Whole merozoite antibody seropositivity thresholds**. (A) IgG (top) and IgM (bottom) levels to the whole merozoite surface for malaria-naive Melbourne adults (n =22), CHMI participants (n = 81) at pre-infection, two weeks post-infection, and EOS, and malaria-exposed residents of Papua New Guinea (n = 5). Seropositivity cutoffs (dashed lines) defined as the mean plus two standard deviations above for reactivity of pre-infection CHMI participants (gold) or malaria-naive Melbourne adults (blue). (B) Seroprevalence of IgG (red) and IgM (blue) to whole merozoites at two weeks post-infection and EOS. Seropositivity cutoffs were defined as the mean plus two standard deviations above for reactivity of pre-infection CHMI participants (left) or malaria-naive Melbourne adults (right).

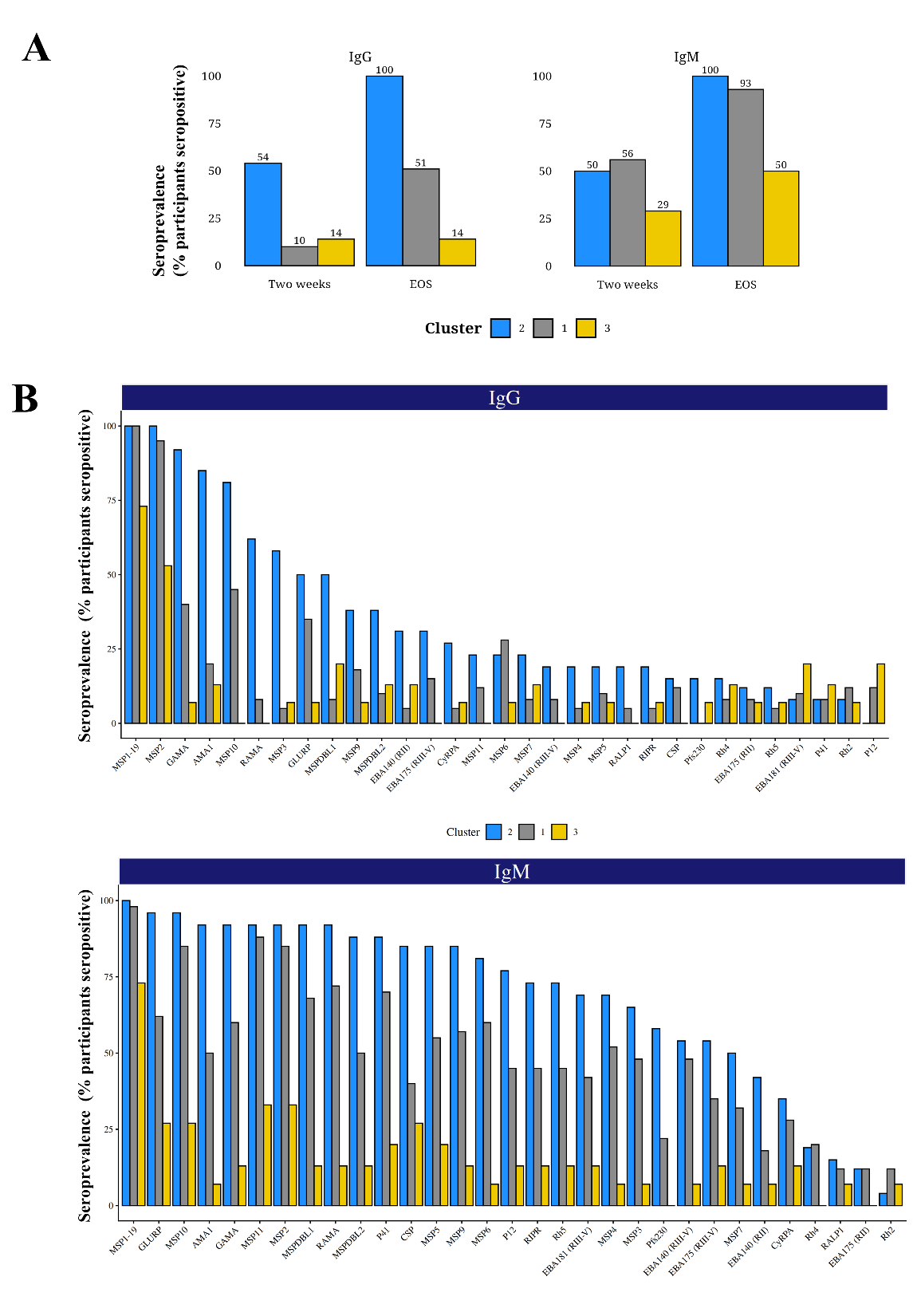

**Supplementary Figure 2**. **Merozoite IgG and IgM breadth across participant clusters**. (A) Seroprevalence of IgG (top) and IgM (bottom) to whole merozoite between participant clusters. (B) Seroprevalence of IgG (top) and IgM (bottom) to P. falciparum antigens with antigens ordered according to IgG seroprevalence for cluster 2.

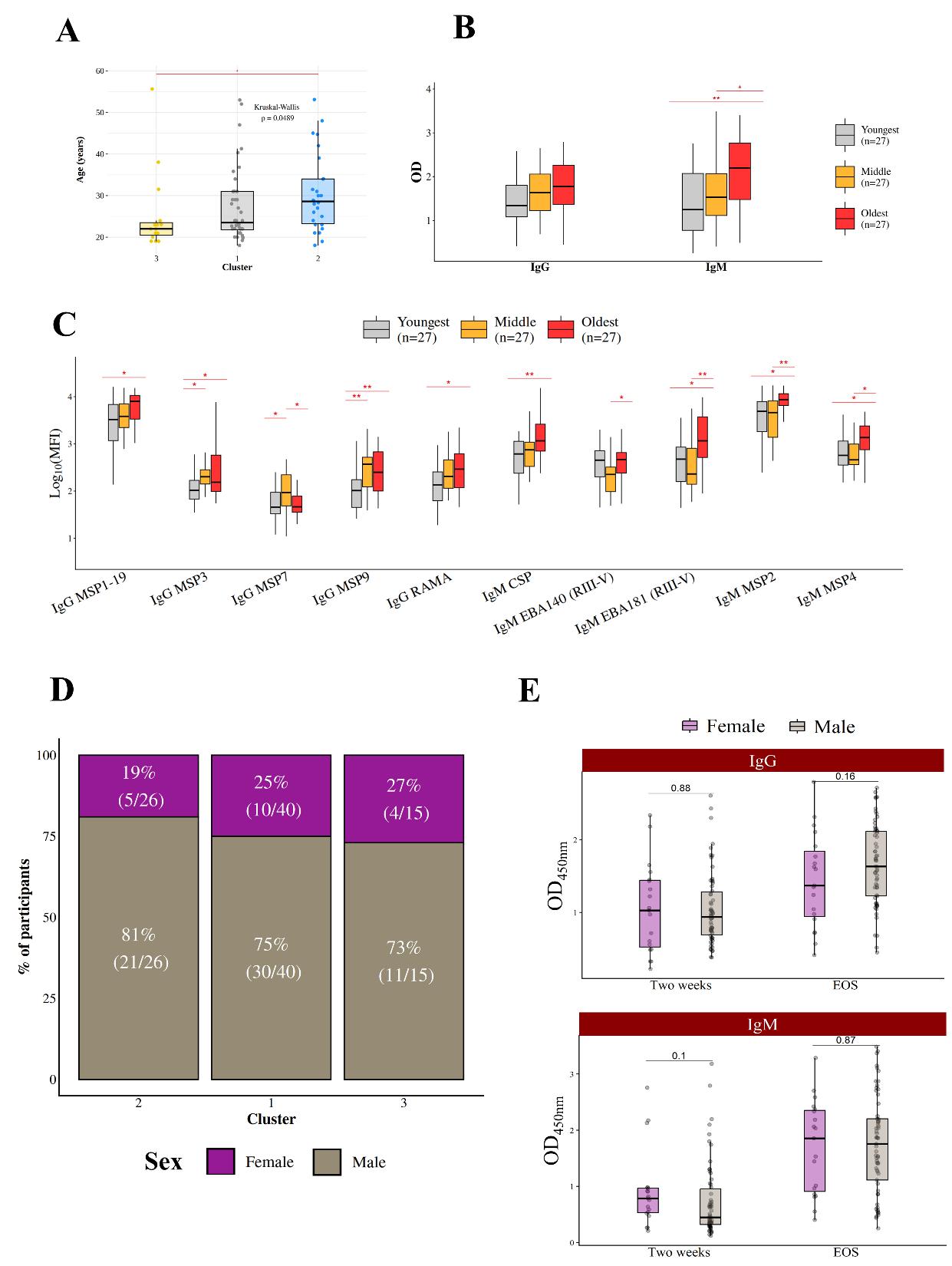

**Supplementary Figure 3. Age associated with more robust merozoite antibody responses following first CHMI.** (A) Age (in years) compared between participant clusters (B-C) Participants were split into even groups according to age tertile, including youngest (18-22 years, n=27), middle (23-29 years, n=27), and oldest (30-55 years, n=27). Age groups were compared using IgG and IgM levels to (B) whole merozoites and (C) P. falciparum antigens. (D) Proportion of participants male (brown) or female (fuscia) across participant clusters. (E) Male and female participants were compared using IgG and IgM levels to whole merozoites at two weeks post-infection and EOS. Group comparisons in A, B, and C performed using Kruskal-Wallis test paired with a Dunn test, whereas comparisons in E performed using a Mann-Whitney U test. Only significant (p < 0.05) comparisons shown.
